# Kinetics and durability of humoral responses to SARS-CoV-2 infection and vaccination

**DOI:** 10.1101/2023.08.26.23294679

**Authors:** Komal Srivastava, Juan Manuel Carreño, Charles Gleason, Brian Monahan, Gagandeep Singh, Anass Abbad, Johnstone Tcheou, Ariel Raskin, Giulio Kleiner, Harm van Bakel, Emilia Mia Sordillo, PARIS Study Group, Florian Krammer, Viviana Simon

**Author notes:** Shared contributions (Komal Srivastava, Juan Manuel Carreño, Charles Gleason, Brian Monahan, Florian Krammer, Viviana Simon). <u>PARIS Study Group</u> (alphabetical order): Hala Alshammary, Angela A. Amoako Dalles Andre, Mahmoud Awawda, Maria C Bermúdez-González, Katherine F. Beach, Dominika Bielak, Gianna Y. Cai, Rachel L. Chernet, Christian Cognigni, Yuexing Chen, Lily Q. Eaker, Emily D. Ferreri, Daniel L. Floda, Miriam Fried, Joshua Z. Hamburger, Denise Jurczyszak, Hyun Min Kang, Neko Lyttle, Julia C. Matthews, Jacob Mauldin, Wanni A. Mendez, Jacob Mischka, Sara Morris, Lubbertus CF Mulder, Ismail Nabeel, Jessica Nardulli, Annika Oostenink, Kayla T. Russo, Ashley-Beathrese Salimbangon, Miti S. Saksena, Amber A. Shin, Levy A. Sominsky, Daniel Stadlbauer, Leeba Sullivan, Morgan Van Kesteren, Temima Yellin, Ania Wajnberg.

## Abstract

We analyzed the kinetics and durability of the humoral responses to severe acute respiratory syndrome coronavirus 2 (SARS-CoV-2) infection and vaccination using >8,000 longitudinal samples collected over a three-year period (April 2020 – April 2023) in the New York City metropolitan area. Upon primary immunization, participants with pre-existing immunity mounted higher antibody responses faster and achieved higher steady-state levels compared to naïve individuals. Antibody durability was characterized by two phases: an initial rapid decay, followed by a phase of stabilization with very slow decay resulting in an individual spike binding antibody steady state. Booster vaccination equalized the differences in antibody levels between participants with and without hybrid immunity, but the antibody titers reached decreased with each successive antigen exposure. Break-through infections increased antibody titers to similar levels as an additional vaccine dose in naïve individuals. Our study provides strong evidence for the fact that SARS-CoV-2 antibody responses are long lasting, with an initial waning phase followed by a stabilization phase.

## Introduction

The emergence of severe acute respiratory syndrome coronavirus 2 (SARS-CoV-2) in late 2019 sparked the global coronavirus disease 2019 (COVID-19) pandemic that is now in its fourth year. Vaccines to mitigate the impact of the pandemic were developed at record speed and have saved millions of lives. However, the emergence of SARS-CoV-2 variants^1^ and waning immunity^2^ have decreased the effectiveness of the vaccines against symptomatic disease^3^. These two issues, emergence of antigenically distinct SARS-CoV-2 variants and waning immunity, are often conflated and used interchangeably but represent two different phenomena^4^. Most vaccines used in North America and Europe are based on lipid nanoparticles (LNP) containing messenger RNA (mRNA) produced by Pfizer/BioNTech (BNT162b2) or Moderna (mRNA-1273), and the common perception now is that mRNA-based vaccine-induced immunity wanes quickly^5^. However, this assumption is mostly based on data from short-term studies that include a very limited number of data points following peak responses^2,5^.

In March of 2020, the densely populated New York metropolitan area was hit with an exponential increase of severe SARS-CoV-2 infections resulting in a staggering number of fatalities and a severely overburdened health care system^6-8^. Due to shortages of personal protective equipment, essential workers in the health care system were at high risk for infection. In response to this crisis, we established (*a*) a specific and sensitive SARS-CoV-2 binding enzyme-linked immunosorbent assay (ELISA) to measure humoral immune responses^9^, and (*b*) an observational longitudinal cohort of health care workers of the Mount Sinai Health System to determine the kinetics of these humoral responses. This study, named *Protection Associated with Rapid Immunity to SARS-CoV-2 (PARIS)^10^*, aims to capture the dynamics of SARS-CoV-2 antibody responses to both infection as well as vaccinations, to determine re-infection rates, and to assess correlates of protection in the context of individual immune histories.

With over 8,000 longitudinal study visits across a single cohort during the first three years of the pandemic, our investigation represents one of the most extensive and in-depth assessments of the longevity of SARS-CoV-2 immune responses to date. Using this longitudinal cohort, we determined the kinetics of antibody responses to spike protein after infections, during the primary immunization series, during monovalent and bivalent booster vaccination as well as during breakthrough infections. Our findings indicate that, in contrast to common perception, COVID-19 mRNA vaccination induces long-lasting antibody responses in humans. The PARIS study also provides insights into the effect of booster vaccination and breakthrough infections on the stability of antibody responses.

## Results

### Study design

PARIS (*Protection Associated with Rapid Immunity to SARS-CoV-2*) is an observational longitudinal study that enrolled 501 adults, mostly healthcare workers (**Table 1**) with or without pre-existing SARS-CoV-2 immunity. The first participants were enrolled in April 2020 when NYC emerged as one of the very early epicenters of the pandemic in the United States. We have conducted over 8,000 study visits with data and biospecimen collection spanning a three-year period (April 2020 to March 2023).

**Table 1:**
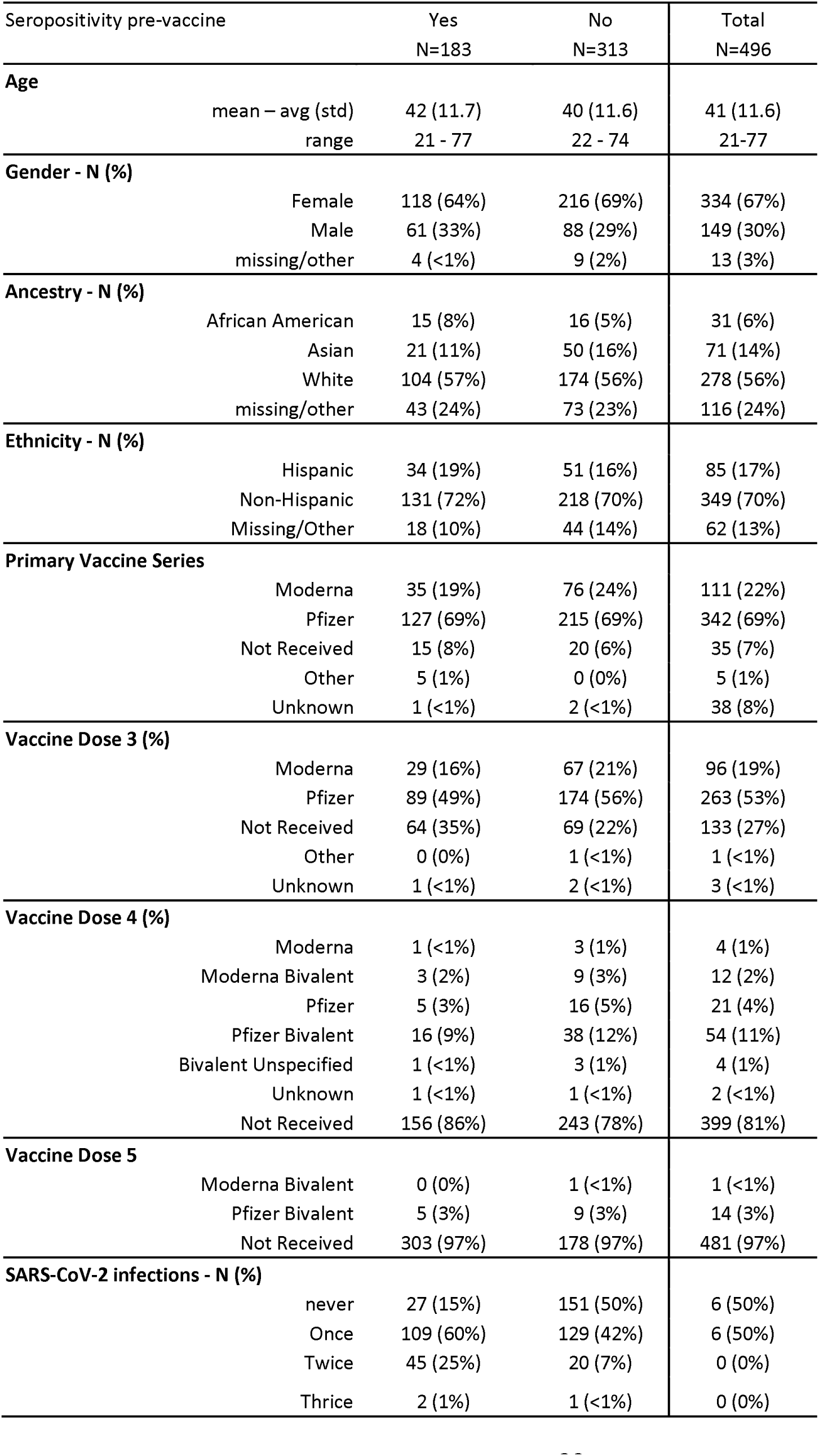
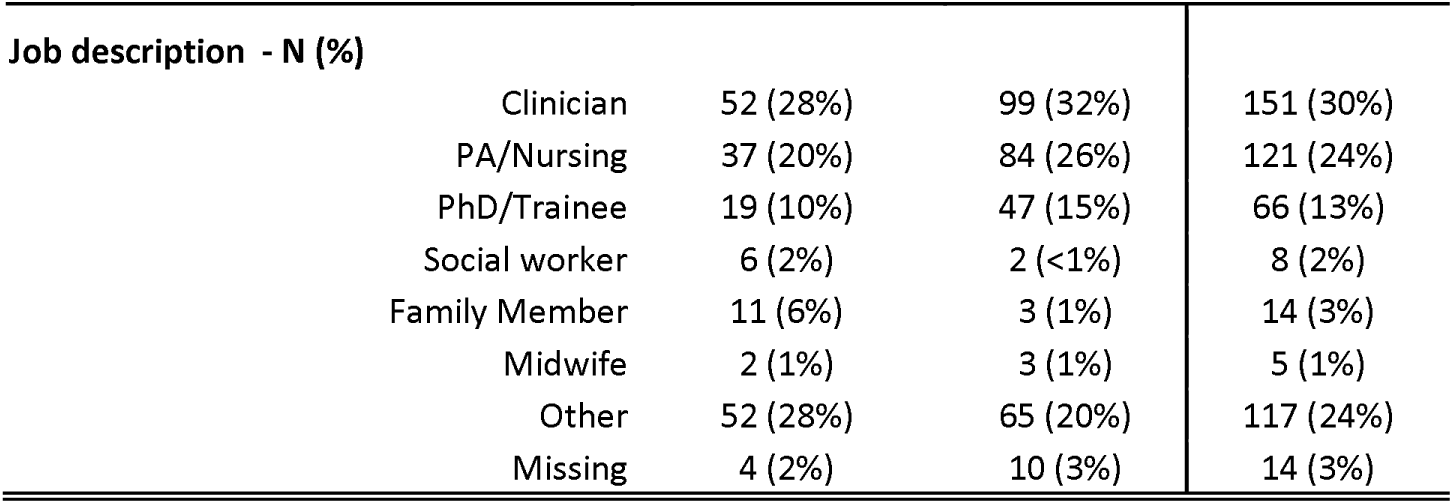
Demographics and Immune histories in PARIS.

Of the participants, 67% were female and 56% self-identified as white. The mean age at study enrollment was 41 years (**Table 1**). At the first study visit, 62% of the participants had no measurable SARS-CoV-2 spike binding antibodies (naive, seronegative). At each study visit we collected data and biospecimen (e.g., blood, saliva). Study visits were scheduled at shorter intervals (2-4 weeks) from study entry through week 8 after enrollment, but the intervals between visits were extended (to approximately 4-8 weeks) for follow-up visits after week 8. Ad hoc study visits were included at short intervals (e.g., weekly) after immune events such as vaccination or infection. Serum samples were used to measure antibodies binding to the ancestral spike protein using an established in-house method that was developed early in the pandemic^9^.

465 of 501 PARIS participants (93%) were vaccinated against SARS-CoV-2 using mRNA vaccines: 342/465 received two doses of Pfizer BNT162b2 and 111/465 received two doses of Moderna mRNA-1273. A small proportion of participants received one dose of the Johnson & Johnson vaccine [Ad26.COV2.S], or two doses of the *CoviShield*™ [AZD1222] vaccine as their primary SARS-CoV-2 immunization regimen (**Table 1**). The number of vaccine doses administered to a given PARIS participant ranged from two to six doses. Briefly, 366/465 (79%) of PARIS participants who completed their primary immunizations subsequently elected to receive a third vaccine dose (“booster”). Of these 366 participants, 97 (27%) received a second booster (4th vaccine dose overall; 27/97 monovalent ancestral booster vaccine versus 70/97 bivalent ancestral/BA.5 vaccine). Of these 97 participants, 15 opted for a third booster (5^th^ vaccine dose overall). Of these 15 participants, one opted for a fourth booster (6^th^ vaccine dose overall). Lastly, 14/27 participants with two monovalent boosters elected to get the bivalent booster.

### Antibody responses to infection and primary immunization

38% of the study participants entered the PARIS study having detectable spike binding IgG antibodies, albeit at highly variable levels (**Figure 1**, area under the curve (AUC) range at baseline: 12.5 to 4,189). Infection-induced anti-spike antibodies showed relatively high stability over approximately 10 months of pre-vaccine follow-up. A detailed analysis of this phase of the study has been reported previously^11^.

**Figure 1:**
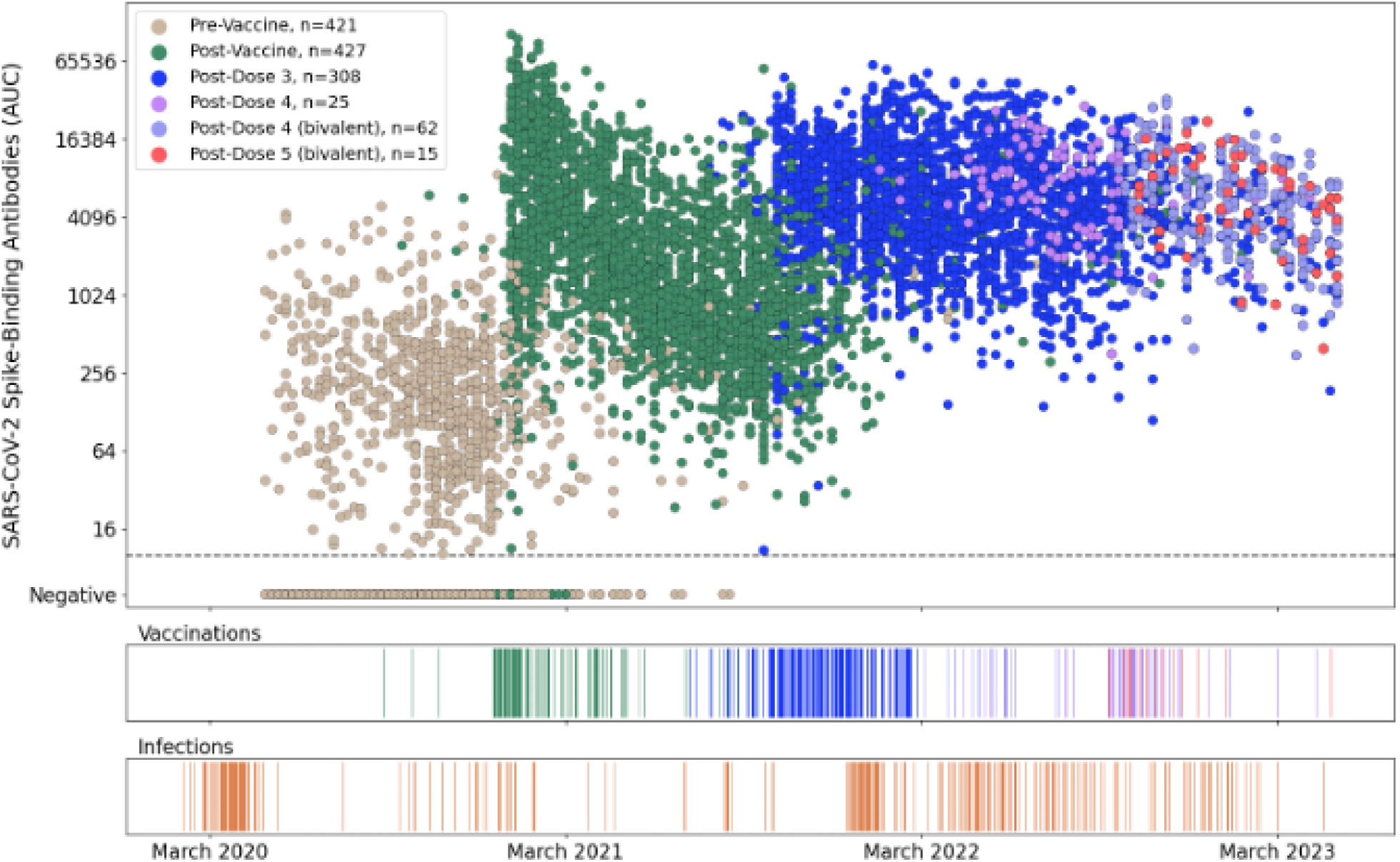
Longitudinal SARS-CoV-2 spike binding antibody levels in 496 PARIS participants over three years. **A**: Each dot represents a distinct study visit at which spike binding antibody levels were measured. Samples are colored by prior vaccination status (2,091 samples pre-vaccination, 3,180 samples post-primary immunization [Dose 1, Dose 2], 2,364 samples post-Dose 3, 110 samples post-monovalent Dose 4, 240 samples post-bivalent dose 4, and 56 samples post-bivalent Dose 5). Each SARS-CoV-2 antibody titer measurement (AUC, area under the curve) is anchored on the Monday of the corresponding week. A small amount of normally distributed noise has been added to the log2-transformed data (σ=0.1), with >95% of transformed values within 15% of the original value to preserve participants’ confidentiality. **B, C**: SARS-CoV-2 infections (**B**) and vaccinations (**C**) events are depicted on the same timeline as the antibody values. Three participants received their primary immunization as part of the Pfizer vaccine trials. The vaccination event colors in the ribbon graph correspond to colors of points after the respective event in the antibody scatterplot shown in Panel **A**.

Vaccinations with mRNA vaccines were available to health care workers in Mid-December 2020 (December 15^th^, 2020, for Pfizer BNT162b2; December 28^th^, 2020, for Moderna mRNA-1273). In naïve individuals, the first dose of the mRNA vaccine induced no to relatively low levels of antibodies (**Suppl. Figure 2A**) but the second vaccine dose, administered 16-31 days after the first (Pfizer BNT162b2: median 21 days [range:16-30]; Moderna mRNA-1273: median 27 days [21-31], increased titers to 5,936 (AUC, peak at 35 days post first dose, **Figure 2A**, **Suppl. Figure 2**). In contrast, individuals with pre-existing immunity induced by infection reached peak titers faster, approximately 10 days after the first dose and achieved significantly higher antibody titers (AUC 19,594, peak at 10 days post first dose (**Figure 2A**, **Suppl. Figure 2**). After completing the primary immunization regimen, participants without pre-existing immunity had > 3-fold lower peak responses compared to participants with pre-existing immunity (p<0.001).

**Figure 2:**
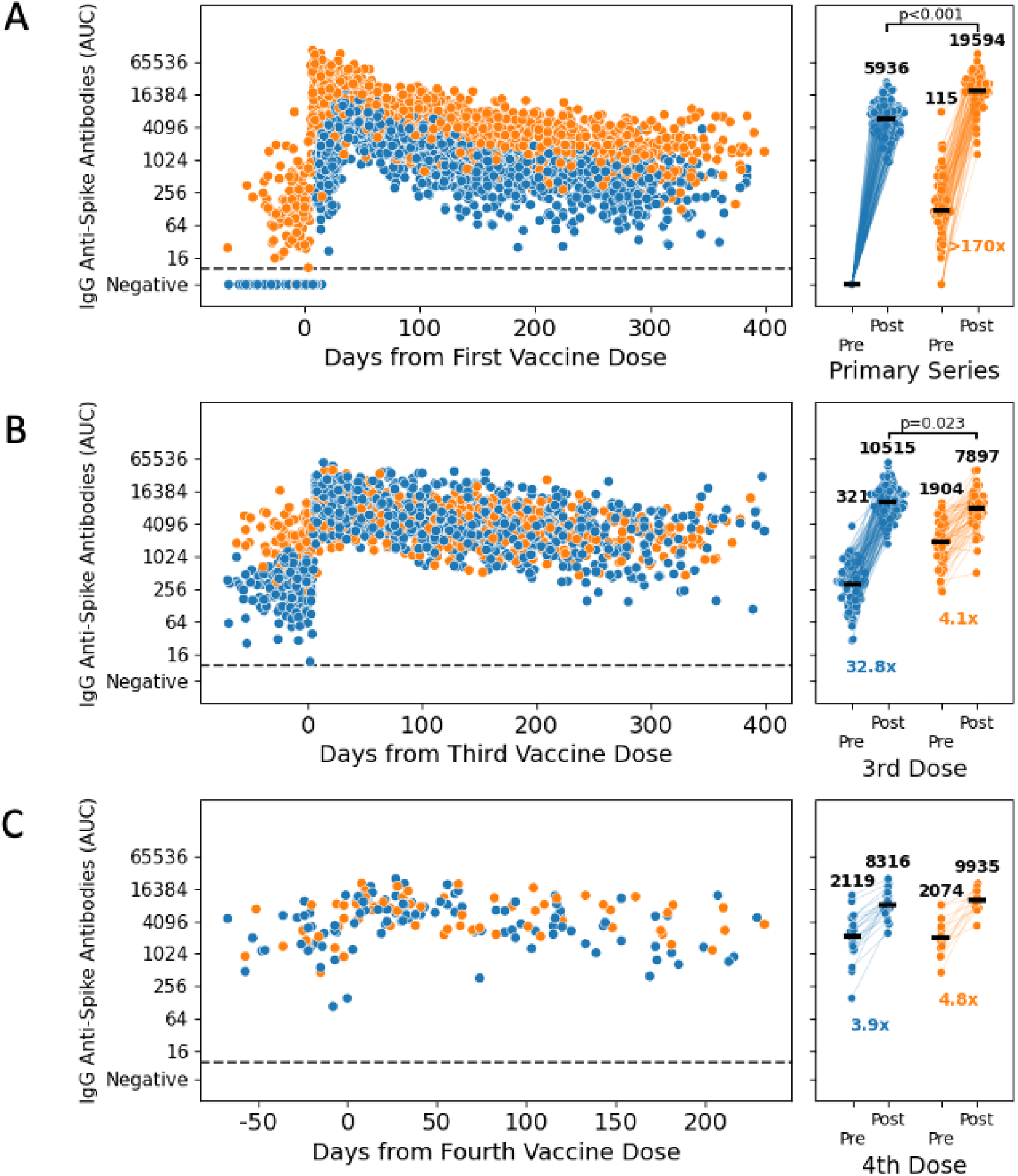
Immunogenicity of the different SARS-CoV-2 vaccine doses. Longitudinal spike binding antibodies measurements (N=4,620) for PARIS participants with (orange) or without (blue) pre-existing SARS-CoV-2 immunity. **A**: Longitudinal antibody follow-up post-vaccination for 179 participants with no prior SARS-CoV-2 infection (blue, 1,671 samples) and 111 participants with a pre-vaccine SARS-CoV-2 infection (orange, 1,083 samples). The right panel has matched pre- and post-vaccine timepoints for 150 previously naïve participants and 92 participants with hybrid immunity. **B**: Longitudinal antibody follow-up pre- and post-third dose for participants with (83 participants, 585 samples) and without (160 participants, 1106 samples) prior SARS-CoV-2 infection. The right panel has matched pre- and post-third dose timepoints for 64 participants with prior infection and 126 participants without hybrid immunity. **C**: Longitudinal antibody follow-up before and after the fourth vaccine dose is shown for 15 participants with SARS-CoV-2 infection prior to fourth dose (67 samples) and 26 participants without hybrid immunity (108 samples). The right panel has matched pre- and post-fourth dose timepoints for 13 participants with prior infection and 21 participants without hybrid immunity. Timepoints post-breakthrough infection were excluded from the analysis. Pre-vaccination timepoints were collected within 10 weeks prior to vaccination while peak post-vaccine timepoints were 1-5 weeks after administration of the vaccine dose (the second dose, in the case of the primary series). The increase in spike-binding antibodies post-vaccination was statistically significant for all recipients (p < 0.0001, Wilcoxon signed rank). Peak antibody levels post-primary vaccination were approximately 3-fold higher in the hybrid immunity group (p < 0.0001, Mann-Whitney U) compared to the vaccine-only immunity group. After third dose, peak antibody levels were modestly elevated in the vaccination-only immunity group compared to those in the hybrid immunity group (p = 0.023, Mann-Whitney U). Peak antibody levels post-dose 4 are comparable across hybrid immunity and vaccination-only groups (no statistically significant difference, Mann-Whitney U).

During the approximately 400 day-long follow-up period, we found an initial steep five-fold drop of antibody titers in vaccinated individuals with and without pre-existing immunity followed by a stabilization phase (**Figure 2A**). Based on a simple rolling geometric mean, post-vaccine data was observed to have two rough “phases” with different rates of decay (**Figure 3A**). This observation resembled a bi-phasic decay that is well-approximated by a two-component three-parameter one-phase exponential decay model framework, prompting us to explore the kinetics of the antibody response in more detail. We fitted a nonlinear mixed-effects (NLME) model to describe the antibody dynamics from two weeks up to one year after the completion of the primary vaccination series. The same model was also fit to antibody dynamics after the third vaccine dose. Specifically, our model has two components: a rapid-decay component with a half-life measured in weeks to months (e.g., antibodies produced by the plasmablast response), and a steady-state component to capture convergence to stable titers during prolonged follow-up (e.g., antibodies produced by long-lived plasma cells) (**Suppl. Figure 3A**).

**Figure 3:**
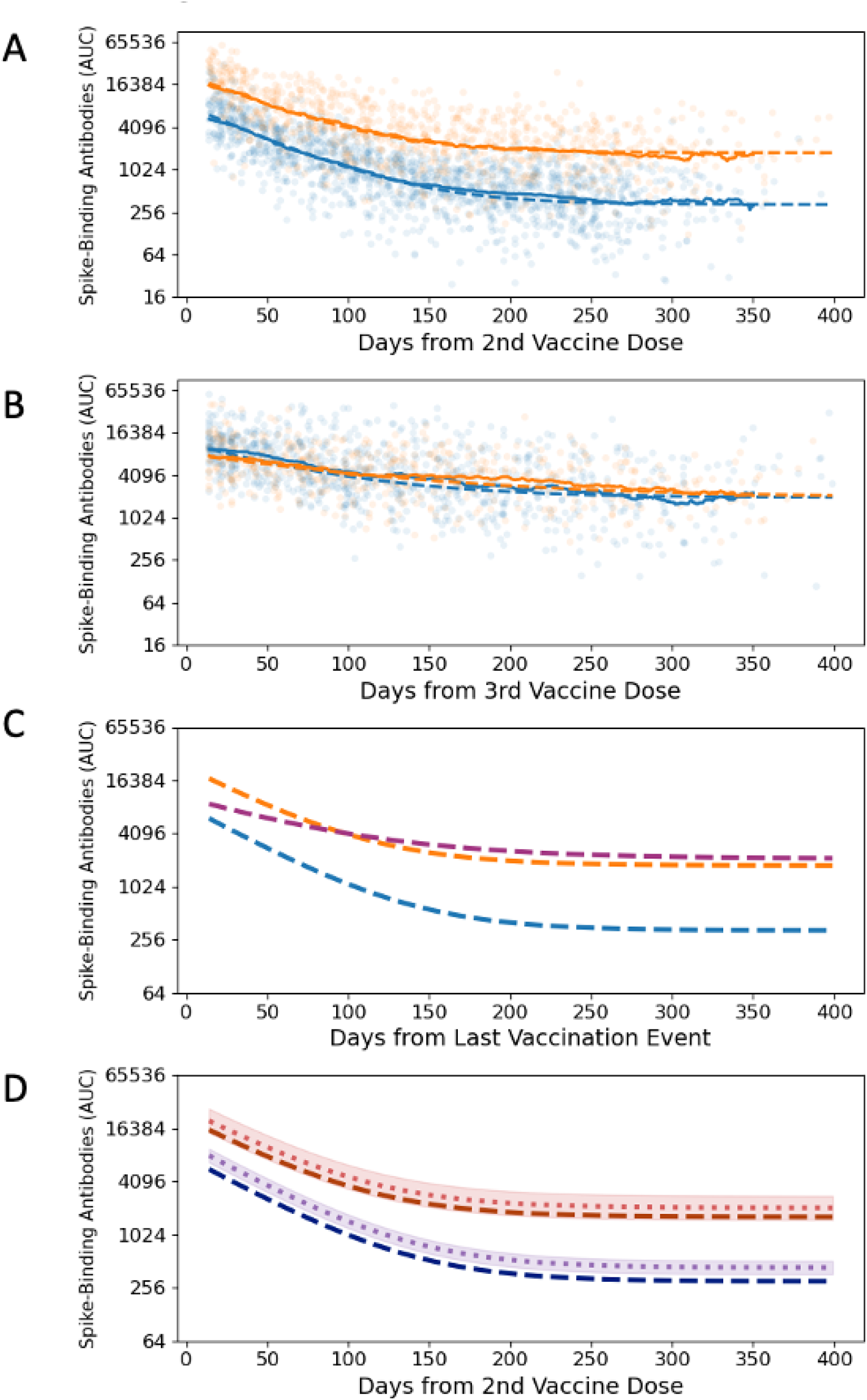
Modelling Antibody kinetics after primary and booster vaccinations. **A**: We generated independent model predictions for the longitudinal post-vaccination antibody levels in 359 participants with (orange dashed line, 126 participants, 850 samples) and without (blue dashed line, 233 participants, 1,443 samples) SARS-CoV-2 infection prior to primary immunization. The rolling 49-day geometric means for each group is shown as solid lines. **B**: We fitted the same model to the longitudinal post-boost antibody levels of 223 participants (80 participants with hybrid immunity, 482 samples; 143 vaccination-only participants, 844 samples). The rolling 49-day geometric means for each group is shown as solid lines. **C**: The model predictions for the dynamics predicted by the post-vaccine model stratified by infection status shown in panel A to a combined post-boost model in dark pink (223 participants, 1,326 samples). **D**: The impact of vaccine type on antibody levels, with the fixed effect due to vaccine type separating model predictions for Pfizer recipients (dark blue and dark red dashed lines) and those for Moderna recipients (purple and red dotted lines). 95% confidence intervals of the fixed effect size are represented by the shaded area. Timepoints after breakthrough infections were excluded from the analysis. 95% confidence intervals are based on a T distribution with standard deviations calculated as part of the model fitting procedure. The vaccine type has a statistically significant effect in participants with no SARS-CoV-2 infection prior to vaccination (67% increase, p < 0.001).

NLME models were fit using the NLMixed procedure in SAS to account for variable longitudinal sampling between participants. A per-participant random effect for each component accounts for the significant variability in both antibody response magnitude and kinetics. Fixed effects due to demographic parameters and vaccine type were fit as multiplicative shifts in the expected geometric mean area under the curve (AUC) across the entire time course. Datasets were stratified by immune status prior to vaccination, with models fit independently to each group. Results for the final models including demographic factors are reported, with independent models fitted for post-vaccine and post-boost antibody dynamics as well as for participants with (“hybrid immunity”) and without (“naïve”) pre-vaccine SARS-CoV-2 infection (**Suppl. Table 1**).

The PARIS NLME model fits closely the observed antibody dynamics post-vaccination, irrespective of prior infection status (**Figure 3A**, **3B**). It also provides the means for a personalized two-component immunity score accounting for an individual’s SARS-CoV-2 immune history reflecting both the magnitude of the peak response and the stability of that response (**Suppl. Figure 3**). These two phases with distinct half-lives distinguish the PARIS model from commonly available models that only consider exponential decay in binding antibodies after vaccination. When comparing our two-phase decay model with a simple exponential decay model, we noted that the latter overestimates the rate of decay in months 6 to 12 post-vaccination in our cohort (**Suppl. Figure 3B**).

For the post-vaccine model (**Figure 3A**), the initial antibody levels measured at 14 days after the second dose are 6,100 AUC in the naïve group (blue) and 17,000 AUC in the hybrid immunity group (2.8-fold higher, in orange). This “peak” response is composed of two components in each case. In the naïve group, the short-lived component has a magnitude of 5,700 AUC (95% Confidence Interval [CI]: 5,200-6,400) with a half-life of 29.6 days (CI: 28.1-31.2) and the stable component has a magnitude of 330 AUC (CI: 290-370). In the hybrid immunity group, the short-lived component has a magnitude of 15,000 AUC (CI: 13,000-18,000) with a half-life of 30.7 days (CI: 28.4-33.5) and the stable component has a magnitude of 1,800 AUC (CI: 1,500-2,100). Because of the larger difference in the stable component, the gap between the hybrid immunity group and the naïve group increases to 4.8-fold at 180 days and 5.4-fold at 360 days after completion of the primary immunization regimen, respectively. Six months after vaccination, the predicted geometric mean antibody titer (GMT) was 445 AUC in the vaccine-only immunity group corresponding to an 18.6-fold reduction relative to the peak antibody levels. In the hybrid immunity group, the predicted GMT was 2,142 corresponding to a 10.7-fold reduction relative to the peak antibody levels. Reductions in antibody levels over the next six months. At 12 months post-vaccination, the predicted GMT was 329 in the vaccine-only immunity group corresponding to only a 1.35-fold reduction relative to antibody levels six months after vaccination. In the hybrid immunity group, the predicted GMT was 1,783 AUC a year after the initial immunization corresponding to a 1.2-fold reduction relative to antibody levels at six months post-vaccination. Thus, although the kinetics of decay in the stable plateau phase are comparable between the two groups, the predicted antibody titer levels for each group differed by more than four-fold (e.g., AUC 329 versus AUC 1,783).

In addition to examining broad patterns in antibody kinetics, we specifically quantified the impact of vaccine type as well as demographic factors including gender, age, race, and ethnicity. Effects were modeled as a constant multiplicative increase or decrease in GMT over the entire time course. Of these, only age and vaccine type were statistically significant at the p < 0.05 level in any group. The effect of vaccine type is illustrated in **Figure 3D** for the post-vaccine model. There is a statistically significant effect in the naïve group, with a 1.43-fold increase in antibody levels (CI: 1.20-1.70) in the Moderna mRNA-1273 vaccine recipients compared to the Pfizer BNT162b2 recipients. There was no statistically significant impact of vaccine type noted in the hybrid immunity group. The effect of age was modest but nonetheless significant in the naïve group: each decade of additional age lived corresponded to a 1.17-fold decrease in antibodies (CI: 1.041-1.187) (**Suppl. Table 1**). In the hybrid immunity group, there was no statistically significant effect.

In summary, based on our data and modeling, antibody titers achieved a steady state seven to nine months after the primary vaccination series. Stable titers were higher for individuals with hybrid immunity as compared naïve vaccinees suggesting induction of long-lived serum antibody titers even after the primary vaccination series. In addition, vaccine type and age had a modest but measurable influence on antibody titer levels in participants without hybrid immunity.

### Antibody responses to booster immunizations

Third dose booster vaccination were authorized for health care workers in September 2021 (September 22^nd^, 2021, for Pfizer BNT162b2 and October 20^th^, 2021, for Moderna mRNA-1273). Booster vaccination resulted in 27-fold increases in peak spike antibody responses in previously naive individuals (**Figure 2B**). In contrast, hybrid immune individuals displayed only a 4-fold increase due to their higher baseline. Four weeks after booster vaccination, the peak titers were similar between previously naïve participants (mean AUC: 10,162) and those with hybrid immunity (mean AUC: 8,001, **Figure 2B**). This stands in stark contrast to the humoral immune responses mounted upon the primary series (**Figure 1**, **Figure 2A**, **Suppl. Fig. 2B**). The third vaccine dose seemed to act as an ‘equalizer’ lifting the antibody titers of participants with vaccine-only immunity for the first time in our longitudinal study to the level of those with infection- and vaccine-induced immunity.

Antibody kinetics post-boost were different compared to kinetics after the primary series (**Figure 3B**). The initial waning of antibodies was slower, and titers stabilized at similar levels to those of the hybrid immune group post-primary vaccination. Importantly, titers settled at comparable levels for both previously naïve and participants with hybrid immunity and stabilized again after approximately seven to nine months. Interestingly, although they stabilized at similar levels, peak levels post boost were moderately but significantly lower than in the hybrid immune group post primary vaccination series.

The fourth vaccine dose did boost serum antibody levels, albeit to a much lower fold-increase compared to prior vaccine doses (vaccine-only 3.9-fold, hybrid immune 4.8-fold) (**Figure 2C**). The peak antibody titers after the second booster vaccine were comparable to the peak titers achieved after the first booster vaccination. Antibody waning kinetics were also very similar to the kinetics observed after administration of the 3^rd^ vaccine dose (**Figure 1**, **Figure 2B**, **Suppl. Fig, 2**). For individuals receiving a third/fourth booster dose (fifth or sixth vaccine dose overall), the data was too sparse for an in-depth analysis, but antibody titers were generally within the same range observed post-3^rd^ and post-4^th^ vaccine doses (**Figure 1**).

We next used our model for the analysis of post boost antibody kinetics. For the post-boost model, antibody levels at 14 days after third dose are 9,500 AUC in the naïve group and 7,400 AUC in the hybrid immunity group (**Figure 3B**). In the naïve group, the short-lived component had a magnitude of 7,500 AUC (CI: 6,500-8,700) with a half-life of 44 days (CI: 39-50) and the stable component has a magnitude of 2,000 AUC (CI: 1500-2500). In the hybrid immunity group, the short-lived component has a magnitude of 5400 UC (CI: 4,300-6,800) with a half-life of 73 days (CI: 60-93) and the stable component has a magnitude of 2,000 AUC (CI: 1,400-2,700). Predicted geometric mean antibody levels 180 days after the beginning of the modeling period (14 days post-boost) are 0.22-fold peak antibody levels in the non-hybrid immunity group and 0.38-fold peak antibody levels in the hybrid immunity group. The rate of decay slows over the next six months, with predicted antibody levels at day 360 of the modeled period are 0.79-fold antibody levels at day 180 in the naïve group and 0.70-fold day 180 antibody levels in the hybrid immunity group. Despite this difference in shape, the geometric mean magnitude of the antibody response is similar across both groups throughout the entire time course studied, with fold-change between groups with and without hybrid immunity ranging from 0.72 to 1.22 and no detectable difference in the magnitudes of the stable component of the model. Because of this, antibody kinetics after boost are well approximated by a single model trained on a combined dataset disregarding prior infection status (**Figure 3B**).

We next compared antibody kinetics after the first booster immunization to those mounted after the primary vaccination (**Figure 3C**). In the model, peak antibody levels after boost are between those for the naïve and hybrid immunity groups post-primary vaccination, and the stable component of the response post-boost is slightly higher in the hybrid immunity group suggesting slower decay. Of note, in the post-boost dataset, the effect due to primary vaccine type follows a pattern like that observed after the primary immunization. Participants without hybrid immunity who received the Moderna mRNA-1273 vaccine for their primary series show a 1.41-fold increase relative to Pfizer recipients (CI: 1.10-1.82). The hybrid immunity group shows no statistically significant difference nor trend due to primary vaccine type. Taken together, our data indicate that booster vaccine doses increase the level at which long term serum antibody responses stabilize.

### Immune responses upon breakthrough infections

While during the early phase of the pandemic, re-infections were rare, breakthrough infections increase with the emergence of variants of concern, especially the Omicron variant. Viral variants of concern started to circulate in the NYC metropolitan area in early 2021 (**Figure 4A**). In our study, we identified breakthrough infections based on participant self-reporting (rapid antigen tests or PCR testing at external facilities), nucleic acid amplification tests and/or an increase in antibody titers (e.g., >4-fold).

**Figure 4:**
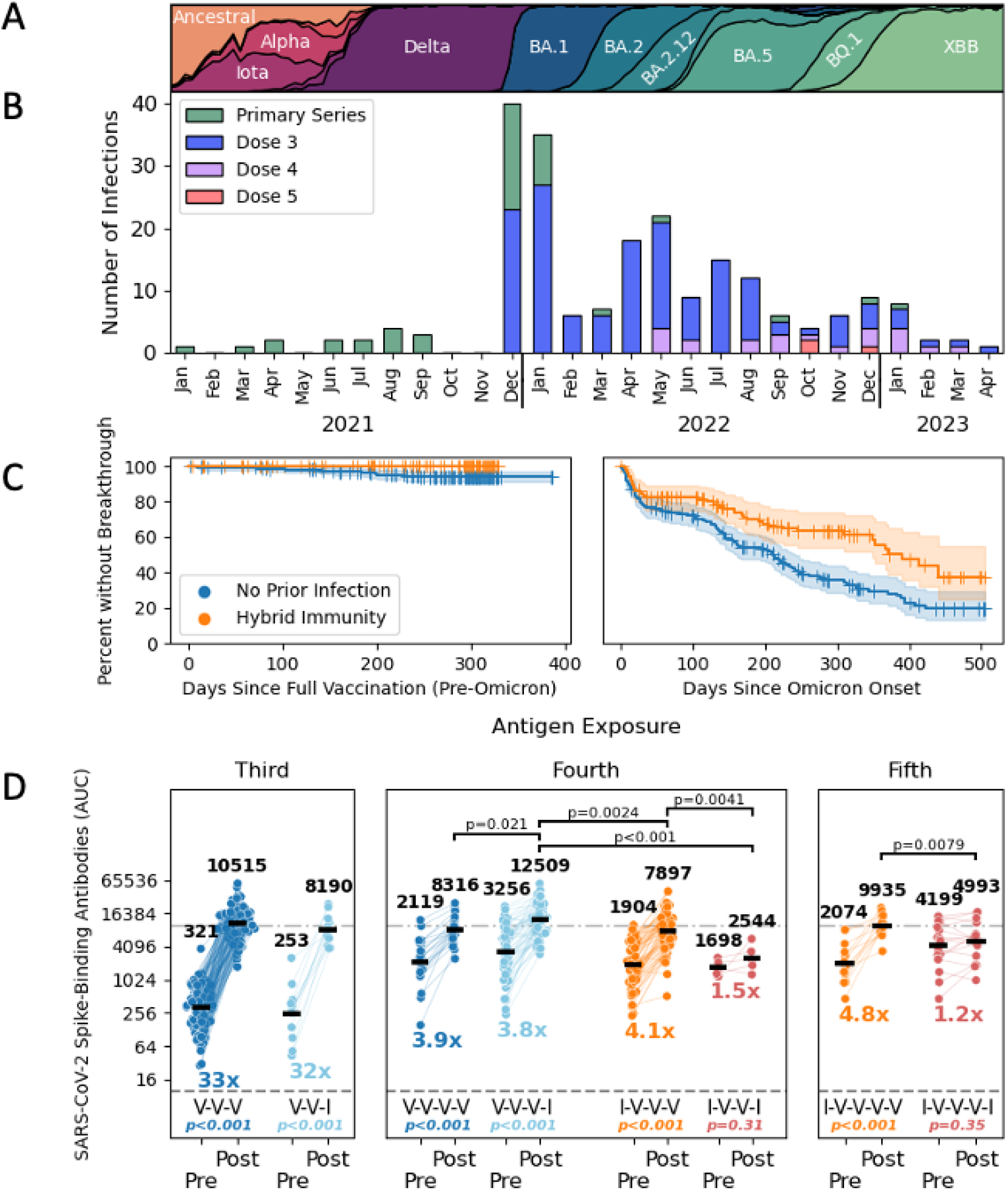
Frequency and impact of breakthrough infections. **A**: The changing SARS-CoV-2 variant landscape in NYC is depicted for the period spanning January 2021 to April 2023.| **B**: The number of breakthrough infections in vaccinated PARIS participants is shown by calendar month (2021-2023). The number of vaccine doses that participants with breakthrough infections received prior to infection are identified by the different colors. **C**: The frequency of breakthrough infections changed after the emergence of Omicron variants in Mid-December 2021. Participants with hybrid immunity were only experiencing breakthrough infections with antigenically diverse Omicron variants (right panel). The differences between vaccinated participants with and without hybrid immunity before (left) and after (right) the appearance of Omicron variants is captured by Kaplan-Meier plots. **D**: The impact of infection and vaccination on spike-binding antibody levels are shown. Vaccination events are shown in blue and orange, with infection events shown in light blue and red. Participants are categorized by number of prior exposures and SARS-CoV-2 infection status prior to initial immunization. Second breakthrough infections are excluded. Third dose (dark blue, n=126) is compared to post-vaccine breakthrough (light blue, n=12). Fourth dose (dark blue, n=21) is paired with the breakthrough post-third dose (light blue, n=54). Third dose in previously infected participants (orange, n=64) is paired with breakthrough re-infection post-vaccine (red, n=5). Finally, antibody titers mounted in response to the fourth dose in previously infected participants (orange, n=13) are compared those mounted after breakthrough re-infections after the third dose (red, n=17). Statistical comparisons within groups (Wilcoxon signed rank test) and fold-change in geometric means are reported below each group. Statistically significant differences in post-exposure antibodies at the p < 0.05 level (Mann-Whitney U test) are reported above.

A total of 225 SARS-CoV-2 infections were recorded among participants over the duration of the PARIS study, with most new infections (214/225) occurring after immunization (at least one vaccine dose administered). We documented three infections after the first vaccine dose and 37 infections after the second vaccine dose but before the booster vaccination (**Figure 4B**). There were 174 breakthrough infections identified in participants who had received at least one booster vaccine dose. The bulk of breakthrough infections were observed after December 20, 2021, when Omicron BA.1 started to spread widely in our community (192/214 new infections). At that point in time, 299 PARIS participants had received a vaccine dose within the prior six months (10/289 second dose of the primary immunization series, 288/289 a booster vaccine dose). 55/299 participants who received a vaccine dose within six months prior to the Omicron wave, experienced a breakthrough infection between December 2021 and February 2022. Nine infections occurred at least 14 days after the participant received a bivalent booster. Of note, one person had three consecutive infections with Omicron variants. Interestingly, before highly antigenically distinct Omicron variants started to circulate in the NYC metropolitan area beginning around mid-December 2021 (**Figure 4A**), most breakthrough infections occurred in vaccinated individuals who were previously naïve, and none occurred in the hybrid immunity group (**Figure 4C**, left panel). After the emergence of the different Omicron lineages, this picture changed, and breakthrough infections also became more common in hybrid immune individuals (**Figure 4C**, right panel), but hybrid immunity continued to have a protective effect as compared to non-hybrid immunity (p=0.00029, log rank test).

We next explored whether breakthrough infections differed from booster vaccination with respect to the increased antibody magnitude. We compared the data for 176 breakthrough infections (at least 14 days after primary vaccination) to the effect observed after receiving an additional vaccine dose (**Figure 4D**). To start, we compared the effect of a booster dose after the primary immunization series (VVV) in participants who had no recorded infection to the change in antibodies in participants who experienced a breakthrough infection after the primary immunization series (VVI). Breakthrough infection in participants who had only had vaccine induced antibodies was the third antigen encounter and increased antibody titers by more than 30-fold, in a manner comparable to an mRNA vaccine booster dose. If the breakthrough infection was the fourth antigen encounter, the fold induction was lower (due to a higher baseline) resulting in an approximately four to five-fold increase which was comparable to a 4^th^ vaccine dose. For the fourth antigen encounter, in the case of a participant whose breakthrough infection was not their first infection and who initially had hybrid immunity (IVVI), however, saw a relatively lower fold change in antibody responses as opposed to participants with hybrid immunity who received a booster dose (IVVV). This difference may be due to the hybrid immunity limiting virus replication during breakthrough infections, leading to less immune stimulation. A similar observation was made when the breakthrough infection as opposed to booster dose was the fifth exposure in participants with hybrid immunity. Especially in formerly naïve individuals, one could argue that breakthrough infections, their first infections, are equivalent to vaccine booster doses in terms of antibody response, while in individuals with hybrid immunity the vaccine has a better effect on boosting systemic antibodies when compared to a second infection.

### Reactogenicity during primary vaccination series and booster doses

In addition to immunogenicity, we analyzed the reactogenicity profiles associated with the different vaccine doses in study participants with and without immunity prior to the primary immunizations. After each vaccine dose, participants were provided a survey to provide self-reported side effects experienced. This analysis builds on preliminary data published in early 2021^12^ which reported that participants with pre-existing immunity experience more systemic side effects after the first vaccine dose compared to naïve participants. We now have data not only on the reactogenicity of the second and third vaccine doses but also longitudinal data on how side effects changed at an individual level.

We collected information regarding local (injection site: pain, erythema, induration, lymphadenopathy) and systemic (e.g., chills, fatigue, fever, headache, arthralgia, myalgia nausea/emesis, pharyngitis) side effects using a survey that allowed participants to self-report the signs and symptoms experienced after each of the first three vaccine doses. 391 PARIS participants provided survey responses regarding symptoms experienced after the 1^st^ vaccine dose, 333 participants provided survey responses to the second dose, and 254 participants responded to the side effects survey after a booster dose. From all survey responses received, we selected the subset of 228 participants (69% female, 70% naïve at primary vaccination) who submitted surveys after each of the first, second and third vaccine doses. Of note, 69% of these respondents received three doses of Pfizer BNT162b2, 21% three doses of Moderna mRNA-1273, and 11% had a mix of vaccine types. Participants without three mRNA vaccine doses were excluded. The distribution of vaccine types was broadly similar across participants with differing immune histories (**Suppl. Table 3**).

Overall, the vaccines were well tolerated with most participant experiencing mild to moderate side effects. Of the participants who reported severe side effects, none required medical attention. 60-72% of the participants who completed all three surveys reported at least one symptom (Dose 1: 60%, Dose 2: 72%, Dose 3: 67%). Across all vaccine doses and independent of infection history, injection site pain is the top reported local side effect and fatigue is the top reported systemic side effect. 11/160 naïve participants and 5/68 participants with hybrid immunity recorded a breakthrough infection prior to the booster dose. Participants with hybrid immunity experienced overall more side effects (**Figure 5**) although the difference in frequency relative to naïve participants became smaller with each additional vaccination. After the first dose, both local and systemic side effects were more common in participants with pre-existing immunity compared to naïve participants (**Figure 5**). After the second dose naïve participants reported a higher incidence of systemic side effects while the frequency of local injection site symptoms remained relatively unchanged. The booster vaccination resulted in less painful injection site reactions in the participants with hybrid immunity (Dose 1/2: 63% vs. Dose 3: 49%) while there was no such difference in the vaccine only immunity group (Dose 2: 52% vs Dose 3: 53%). Both groups also reported lymphadenopathy (e.g., axillary) to occur more frequently (7-11%) while fatigue remained unchanged (43-44%). The incidence of chills jumped from 6% to 28% in the naïve group while participants with hybrid immunity only experienced a two-fold increase (19% to 32%) upon booster vaccination. **Suppl. Table 4** provides a summary of the frequency of side effects. When analyzing the data, age had no observable trend for the incidence or severity of reported vaccine reactogenicity. Female participants reported more side effects in most categories when compared to male participants, but this could also be a bias from the side effects respondents’ group as most participants are female. In the naïve participant subgroup, Moderna mRNA-1273 vaccinees experienced more chills, nausea, arthralgia, fever, headache, erythema and induration at the injection site after the second and third dose of the vaccine compared to Pfizer vaccinees. The median duration between 2^nd^ and 3^rd^ vaccine doses was 272 days (range: 155-400).

**Fig. 5:**
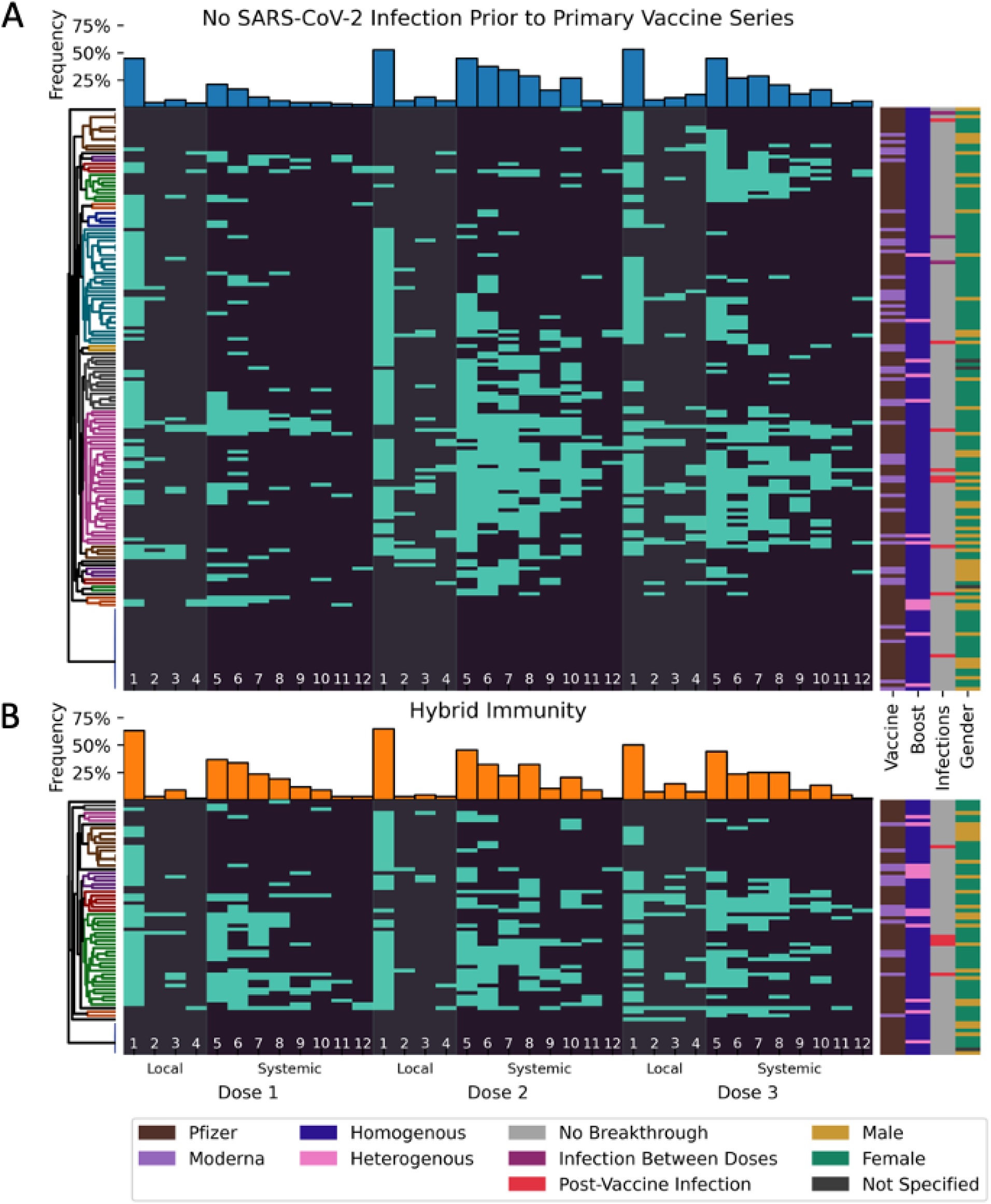
Reactogenicity reported after sequential SARS-CoV-2 accination (Dose 1, 2 and 3) **A, B**: Each line represents the post vaccine side effects reported by the same participant. Data shown is based on 684 surveys completed by 228 participants (160 initially naïve, [70%, **Panel A**] and 68 with hybrid immunity, [30%, **Panel B**]) after the first, second and third vaccine dose. The order of the symptoms depicted are divided into side effects at the local injection site (pain [1], erythema [2], induration [3], lymphadenopathy [4], grey bands) and systemic in nature (fatigue [5], headache [6], myalgia [7], chills [8], arthralgia [9], Fever [10], nausea/emesis [11], pharyngitis [12], dark grey background). Participants are split into groups based on pre-vaccine infection status and ordered by UPGMA clustering based on the Jaccard metric with optimal leaf ordering (trees shown on the left, colored clusters contain all nodes at distance less than 0.7). The presence of a symptom is indicated by a light blue bar. Bar plots for the overall frequency of each symptom are shown above the longitudinal data for each group. Vaccine type, boost type, infection status, and gender for each participant are annotated on the right.

To better capture how vaccine reactogenicity changes between the three vaccine doses and visualize symptom clusters, we performed Unweighted Pair Group Method with Arithmetic Mean (UPGMA) clustering based on Jaccard distance with optimal leaf ordering (**Figure 5**). This ordering showed that, while overall frequencies of side effects appeared comparable between Dose 2 and 3, this does not hold true at the individual participant level. Indeed, some participants with vaccine induced immunity experienced little or no symptoms after the second dose but reported several systemic side effects after the third dose while others had far fewer side effects after the booster vaccination compared to the second dose.

Taken together, these real-world data indicate that pre-existing immunity modulates the reactogenicity with additional SARS-CoV-2 vaccine doses resulting in more pronounced side effects. Booster vaccine doses produced slightly fewer systemic side effects relative to the second vaccine dose in naïve participants while the opposite is true for those with hybrid immunity.

## Discussion

The PARIS study^10^ aimed to investigate the durability of immune responses mounted to SARS-CoV-2 infection and vaccination. Indeed, the SARS-CoV-2 pandemic provided unprecedented challenges but also opportunities to address fundamental questions in human immunology. A large proportion of the human population was exposed — for the first time and within a very limited time window of the pandemic — to a new respiratory viral pathogen and to a series of sequential immunizations that included an antigen to which the whole population was naive.

We used our extensive longitudinal antibody data with frequent sampling to address several important open questions about SARS-CoV-2 antibody-based immunity. It is often assumed that immunity after vaccination with mRNA vaccines wanes quickly. In some cases, this is based on the observation that vaccine effectiveness decreases over time^2^. However, declining vaccination effectiveness is impacted not just by waning immune responses, but also by virus evolution^13^. In other cases, the observation window is limited to the peak response and a brief period, thereafter, producing a linear decay model that does not accurately reflect B-cell biology. Finally, many studies analyze receptor binding domain-focused neutralizing antibodies only but ignore the large number of other epitopes present on the spike protein. Our study design allows us to follow individuals longitudinally from the onset of the pandemic, through primary immunization series and now after booster immunizations providing an invaluable opportunity to comprehensively analyze long-term kinetics of antibody-based immunity (**Figure 1**). We found that anti-spike antibody kinetics after the primary immunization series follow a pattern that would be expected from basic B-cell biology. After primary vaccination, serum antibodies produced by plasmablasts reach a high peak^14^. Plasmablast responses are the first line of defense when it comes to B-cells^14^. These cells proliferate quickly, circulate in the periphery, and produce large amounts of antibodies – but typically disappear within two weeks^15,16^. The antibodies they produce have a longer half-life (e.g., IgG1 has a half-life of approximately four weeks^17^), but eventually also wane over the period of weeks to months. This is consistent with the 28-to-34-day half-life of the short-lived component of our PARIS model following the primary vaccination series. However, serum antibody responses then stabilized after the initial few months. The more stable long-term serum antibody response is likely produced by long-lived plasma cells which have in the meantime migrated from lymph nodes into the bone marrow^18^. Again, the observations in our cohort basically follow textbook B-cell biology. Interestingly, waning kinetics were slower after the booster doses and the level at which titers stabilized became higher after each booster dose. We noted a clear “ceiling” effect with respect to the overall peak antibody titers reached. While initial titers after the primary vaccination series in previously infected individuals could reach almost 20,000 AUC, overall, the titers in general seemed to peak at approximately 10,000 AUC. This ceiling seemed to slowly become lower with repeated exposures and booster doses (**Figure 2**).

A key contribution of our study is the development of a robust model suitable for dissecting the biological factors contributing to the durability of SARS-CoV-2 immune responses (**Figure 3**). Leveraging our extensive longitudinal antibody data, we could identify patterns and associations that shed light on the mechanisms underpinning long-term antibody protection. Our model allows us to interrogate the determinants of antibody dynamics, including prior SARS-CoV-2 infection and vaccine type as well as age. We found that individuals with initial infections followed by vaccination had antibody decay kinetics similar to individuals who were naïve before the primary vaccination series, however their peak titers were higher^12^, and so was the titer at which their long-term serum antibodies eventually settled. Interestingly, the booster dose acted as an equalizer, bringing the serum antibody to similar levels in the group who was initially infected and the group that was initially naïve. Other factors that influenced antibody titers, including long-term stable titers, were age and vaccine type. As observed in other studies that stated superiority of Moderna mRNA-1273 over Pfizer BNT162b2 in terms of induced immune responses or protection^19,20^, we saw that the Moderna mRNA-1273 vaccine induced approximately 1.3-fold higher titers than the Pfizer BNT162b2 vaccine, although differences were only significant in initially naïve individuals.

Our study also allowed us to determine infection rates in our cohort. We found that during the pre-Omicron era very few breakthrough infections occurred and only in the previously naïve group (**Figure 4**). During this time not a single breakthrough infection was detected in the hybrid immune group over the course of 11 months. This changed significantly during the Omicron era when the majority of our participants experienced breakthrough infections over the course of 18 months. This observation makes sense since Omicron has a strong escape phenotype and its spike mutations undermine neutralizing antibodies induced by the ancestral or earlier variant spikes^1^. However, a notable protective effect remained in the hybrid immune group as compared to initially naïve individuals (**Figure 4C**). We also assessed the immune responses to breakthrough infections. When comparing the number of antigen exposures, we found that a breakthrough infection after the primary vaccination series in previously naïve individuals induced antibody titers comparable a third dose of vaccine (booster dose). We found a similar pattern for the fourth exposure (4^th^ vaccine dose versus breakthrough infection after the 3^rd^ vaccine dose). This suggests that these initial breakthrough infections in vaccinated but previously naïve individuals do in fact act similarly to a booster dose in terms of inducing antibody responses. The picture changed in individuals who had an initial infection followed by two or three vaccinations. When comparing breakthrough infection versus one additional vaccination in those highly exposed individuals we saw that vaccination still robustly boosted serum antibody titers but breakthrough infection did not, at least not as robustly as vaccination. We speculate that virus replication in these individuals was highly restricted due to their strong pre-existing immunity and that therefore the presence of less antigen leads to a lower induction of immunity.

Our longitudinal study setting also allowed us to investigate vaccine reactogenicity as it was experienced after the first, second and third vaccine dose in a longitudinal manner at individual participant level. These data are of great relevance since reactogenicity can increase vaccine hesitancy and reduce uptake. In PARIS, SARS-CoV-2 mRNA vaccines were generally well tolerated with mild to modest side effects occurring in two thirds of the vaccinees. As reported by us and others^12,21^, both local and systemic side effects were more common in individuals with hybrid immunity (**Figure 5**). Our participant level analysis of vaccine reactogenicity reveal the landscape of reactogenicity with several symptom clusters being apparent in the participant group with no infection prior to vaccination. In future studies, we plan on analyzing whether certain pattern of vaccine side effects predict the level of immunogenicity. These real-world data on side effects associated with sequential vaccine doses is important to provide guidance for individuals concerned regarding future immunizations.

Our study has several limitations. We were unable to analyze mucosal immune responses which are especially interesting during breakthrough infections. We and other have previously shown that secretory IgA antibodies are not induced or boosted by mRNA vaccination in individuals who were previously naïve but that titers increase in hybrid immune individuals after their first vaccination^22,23^. We plan, therefore, to conduct similar studies at additional time points in our cohort in the future. Additionally, because our purpose was to analyze serum antibody kinetics, the current study did not measure neutralizing antibody responses since binding antibodies contain all antibodies to the spike and not just antibodies to certain neutralizing epitopes. Similarly, the current study focused on antibodies to the full-length spike rather than RBD. However, since we have observed differential antibody kinetics for spike and RBD^24^ (which is part of the spike), we plan to measure RBD antibodies in the future. We plan to assess antibody avidity in our longitudinal cohort using different methods as previously described^25^. Of note, binding antibodies have been tied to protection from severe COVID-19 outcomes in the absence of neutralizing antibodies in a recent study^26^. Finally, the current study examines reactivity to ancestral spike, and not to later variant spikes or nucleoprotein. Assays utilizing the ancestral spike also detect antibodies induced by variant breakthrough infection and existing bivalent vaccines. Indeed, it has been shown that no to little specific antibodies are generated to new spikes and most of the response is cross-reactive^27,28^ in a manner consistent with ‘backboosting^29^.

In summary, we show that antibody responses to SARS-CoV-2 mRNA vaccination follow a classical biphasic decay with a rapid waning phase initially followed by a transition into a stabilization phase after 7-9 months. We also show that hybrid immunity showed better protection against breakthrough infection in both the pre-Omicron but also the post-Omicron era and we show that breakthrough infections in vaccinated, previously naïve individuals have an effect like a booster dose. These data suggest that COVID-19 mRNA vaccination does induce long lasting spike-specific antibodies consistent with B-cell biology.

## Data Availability

All data produced in the present study are available upon reasonable request to the authors after publication

## Acknowledgements

We thank all the participants of our longitudinal PARIS study for their generous and continued support of our COVID-19 research programs.

This work was funded in part by the Centers of Excellence for Influenza Research and Surveillance (CEIRS, contract # HHSN272201400008C), the Centers of Excellence for Influenza Research and Response (CEIRR, contract # 75N93021C00014), by the Collaborative Influenza Vaccine Innovation Centers (CIVICs contract # 75N93019C00051), by philanthropic donations and by institutional funds including the Mount Sinai Center for Vaccine Research and Pandemic Preparedness.

This effort was also in part supported by the Serological Sciences Network (SeroNet) in part with Federal funds from the National Cancer Institute, National Institutes of Health, under Contract No. 75N91021F00001 via 21X092F1 Mod 01. The content of this publication does not necessarily reflect the views or policies of the Department of Health and Human Services, nor does mention of trade names, commercial products or organizations imply endorsement by the U.S. Government.

## Table of content for the Supplemental Online materials

- Online Methods
- Supplemental Tables S1 to S3
- Supplemental Figures S1 to S4

## Online Methods

### Study Information

In Spring 2020, most of New York City was shut down except for essential personnel needed to fight the SARS-CoV-2 pandemic. Personal Protective Equipment was in high demand and needed to be distributed to essential workers providing high risk services. Health care workers performing a wide array of different jobs (e.g., from housekeeping, students, nurses to medical doctors) fell into this category.

The observational longitudinal PARIS Study (Protection Associated with Rapid Immune response to SARS-CoV-2) enrolled adult individuals who worked at the Mount Sinai Health System (MSHS) in New York City in April 2020. Adult family members of MSHS employees were also eligible for participation. The study protocol IRB-20-03374 / STUDY-20-00442 was approved by the Institutional Review Board of the Mount Sinai Hospital. All study participants signed written consent forms prior to providing data or biospecimen.

In the first two months of the study, participants were asked to provide saliva samples weekly and to provide a blood sample every two weeks. After the two-month study visit, participants returned monthly for a period of one year, thereafter study visit frequency varied between one and two months. Nasopharyngeal or ante-near swabs were collected when participants reported symptoms suggestive of a respiratory infection. Ad hoc study visits were performed in these situations as well as after vaccination. If participants reported testing positive for a viral respiratory infection, details about the diagnostic test performed, symptom onset and disease severity were collected.

A total of 501 health care workers were enrolled in PARIS between April 2020 and August 2021. Prior to the SARS-CoV-2 vaccine rollout in December of 2020 412/501 participants had been enrolled. Based on serological status at baseline prior to vaccination, 38% of participants had SARS-CoV-2 spike binding IgG antibodies above the limit of detection of an in-house ELISA test. One single participant reported a SARS-CoV-2 infection prior to enrollment into the study but was seronegative at baseline.

Of the 501 participants enrolled, one (0.2%) has received four mRNA booster doses (total vaccine doses: 6), 13 (2.6%) have received three mRNA booster doses (total vaccine doses: 5), 83 (16.6%) have received two mRNA booster doses (total vaccine doses: 4) and 269 (53.7%) received one booster vaccine dose. 465 participants (93% of the total cohort) have been vaccinated with the remainder of the participants having withdrawn or being lost to follow up since immunization became mandatory for health care workers. Indeed, 36 participants withdrew from the study and their vaccination status is unknown. Of the 465 participants who were vaccinated, 99 (21.3%) elected not to receive any booster vaccine dose. Over the three years of the PARIS study, we have documented over 8,000 study visits collecting blood, saliva and/or nasal swabs.

### Vaccine reactogenicity

Qualitative survey data capturing general health questions, exposures, and changes to perceived risk to SARS-CoV-2 infection were captured monthly using a custom build REDCap database. When participants received a SARS-CoV-2 vaccination, a survey to collected information about side effects was sent out after the 1^st^ and 2^nd^ dose in the primary immunization series as well as after the 1^st^ booster vaccine dose.

After vaccination, surveys were sent to participants to provide data on side effects experienced after dose 1, 2 and 3. The questions for the survey were designed to capture diversity of possible effects broken into two categories: systemic effects and local injection site effects. The symptoms recorded (**Suppl. Table 2**) included side effects reported in publications from Pfizer and Moderna about their respective vaccines^30,31^.

Dates by which vaccines became available to health care workers were as follows: Pfizer BioNTech: 12/15/2020 (initial) and 09/22/2021 (booster); Moderna: 12/28/2022 (initial), 10/20/21 (booster). Bivalent boosters (both Pfizer/BioNTech, Moderna) were distributed starting September 2022. Three participants received vaccine in the Pfizer Phase 3 vaccine trial.

The surveys for the first two doses were released early in 2021 and the booster survey was launched in the fall of 2021. 245 participants completed all three surveys. Participants who did not receive mRNA vaccination as their primary immunization or booster dose were excluded from the analysis as well as participants with known immunomodulatory comorbidities leaving a total complete survey response from 228 participants. For each indicated side effect, branching logic questions were designed to ask about the duration of the side effect and perceived severity. For fever and nausea/vomiting, quantitative ranges were asked. For fever, we asked about the highest temperature known. For nausea/vomiting we asked about the number of times a day the participant felt ill and vomited. The other side effects, if indicated as severe, had follow-up questions to ask if medical care was required. None of the participants who reported a severe side effect required hospitalization for treatment. All survey data was captured using REDCap database system. Severity scoring was tabulated in a REDCap report output (not reported = 0, mild = 1, moderate = 2, severe = 3) per effect per vaccine dose. The dataset for all three doses was combined and subgroups based on infection history pre-vaccine, vaccine type, and gender were created. Percent totals of reported incidence and severity were calculated per subgroup and then compared to understand differences within the PARIS cohorts and observe trends between groups across doses.

### Enzyme-linked immunosorbent assay (ELISA)

Antibody titers were measured using a two-step well-established ELISA method^9,32^ in which serum samples are screened at a single dilution (1:50) for IgG against the recombinant receptor binding domain (RBD) of the spike protein from SARS-CoV-2 (Wuhan-Hu-1), followed by detection of antibodies against the full-length spike protein (also Wuhan-Hu-1). In the first step, serum samples were screened in a high-throughput assay using the recombinant RBD protein. In brief, 96-well microtitre plates (Thermo Fisher) were coated with 50 μl recombinant RBD protein at a concentration of 2 μg/ml overnight at 4 °C. The next day, the plates were washed three times with phosphate-buffered saline (PBS; Gibco) supplemented with 0.1% Tween-20 (PBS-T; Fisher Scientific) using an automatic plate washer (BioTek). The plates were blocked with 200 μl blocking solution consisting of PBS-T with 3% (w/v) milk powder (American Bio) and incubated for 1 h at room temperature. Serum samples were heated at 56 °C for 1Lh before use to reduce the risk from any potential residual virus in the serum. The blocking solution was taken off the plates and 100 μl of the serum samples diluted 1:50 in PBS-T containing 1% (w/v) milk powder was added to the respective wells of the microtitre plates. After 2 h, the plates were washed three times with PBS-T and 50 μl anti-human IgG (Fab-specific) horseradish peroxidase antibody (produced in goat; Sigma, A0293) diluted 1:3,000 in PBS-T containing 1% milk powder was added to all wells and incubated for 1 h at room temperature. The microtitre plates were washed three times with PBS-T and 100 μl SigmaFast o-phenylenediamine dihydrochloride (Sigma) was added to all wells. The reaction was stopped after 10 min with 50 μl per well 3 M hydrochloric acid (Thermo Fisher) and the plates were read at a wavelength of 490 nm with a plate reader (BioTek). Serum samples that exceeded an optical density at 490 nm (OD490) cut-off value of 0.15 were categorized as presumptive positive and were tested in a second step in confirmatory ELISAs using the full-length, recombinant spike protein. To perform the confirmatory ELISAs, the plates were coated and blocked as described above except full-length spike protein at a concentration of 2 μg/ml was added to the plates. After 1 h, the blocking solution was removed, presumptive positive serum samples that were serially diluted (from 1:80/1:100 to 204,800) in 1% milk prepared in PBS-T were added and the plates were incubated for 2 h at room temperature. The remainder of the assay was performed as described above. The cut-off value was set to an OD_490_ value of 0.15 and true-positive samples were defined as samples that exceeded an OD_490_ value of 0.15 at a 1:80/1:100 serum dilution. The end-point titer was calculated and defined as the last dilution before the signal dropped below an OD_490_ of 0.15. The area under the curve (AUC) values were calculated and plotted using Prism 9 software (GraphPad). AUC values >10 were plotted as above the limited of detection in the figures included in this manuscript.

### SARS-CoV-2 variants

Genomic data on the viral variants circulating within the New York City metropolitan area were obtained through the New York City Department of Health and Mental Hygiene (DOHMH) web portal. The DOHMH maintains a publicly available database updated regularly since the beginning of the pandemic. SARS-CoV-2 variant tracking within the database begins on January 1^st^, 2021, and was updated weekly until May 18^th^, 2023 (when the COVID-19 emergency was declared over in the US). Data on specific viral variants was only available for cases sequenced by and/or shared with the NYC DOHMH. Sequences were not available for all the SARS-CoV-2 positive test cases reported to DOHMH. Certain variants within the NYC data were collapsed into single categories due to the limited size of variant. The following variants were combined: XBB and XBB 1.5; BQ1 and BQ 1.1, Omicron BF.7, BA.4 and BA.4.6; B.1.427 and B.1.429 (Epsilon), and finally the B.1.526 lineages with and without E484K (Iota).

### Statistical Analysis

Statistical testing for paired samples was done using the Wilcoxon signed rank test, with unpaired samples compared using the Mann-Whitney U test. The protectiveness of hybrid immunity was evaluated using a log rank test. UPGMA clustering for Figure 5 and all statistical tests were run using Scipy (version 1.11.1) in Python 3.9.15. Curve fitting for **Supplemental Figure 3** was done using the statsmodels package (version 0.13.2). All figures were rendered using Matplotlib (3.7.1) and Seaborn (0.12.2). The antibody kinetic model was fit using the NLMixed procedure in SAS9.4, using the trust region optimization technique and the adaptive gaussian quadrature integration method, assuming a normal distribution for the random effects and the dependent variable.

### Non-Linear Mixed models for spike binding antibodies

Data were selected for the post-vaccine model under the following criteria: Timepoints with a discrepancy between the results generated by the research grade SARS-CoV-2 spike-binding ELISA and the Kantaro spike-binding assay performed in the CLIA certified Pathology laboratories of the Mount Sinai Hospital in the top or bottom percentile over the entire dataset were excluded. Participants who did not receive two doses of an mRNA vaccine as well as participants with unknown SARS-CoV-2 infection status prior to immunization were also excluded. Similarly, data from participants without at least two valid timepoints 14 days post-dose 2 were excluded. Samples obtained after subsequent vaccine doses or breakthrough infections were censored.

Data were selected for the post-boost model using the following criteria: Participants excluded from the post-vaccine model dataset were also excluded from the post-boost dataset. Participants with a breakthrough infection pre-third dose were excluded. Samples after fourth doses of the vaccine or breakthrough infections were censored. Participants without a minimum of two valid time points at least 14 days post-dose 3 were excluded.

Antibody decay has been observed in interventional settings where external antibody treatments were administered. Unlike clinical treatments, natural immunity has multiple sources of antibody production. Since there are multiple sources of antibody production naturally within the body, single component exponential decay models are insufficient as they will over-predict the change in decay rates. The model used here has two components which are assumed to have distinct decay rates. When reporting aggregate antibody data, the geometric mean is generally used due to a high variability within and among participants (across multiple orders of magnitudes in the datasets used here). By fitting the model directly within the log-transformed space, the output curve directly estimates the geometric mean rather than the arithmetic mean.

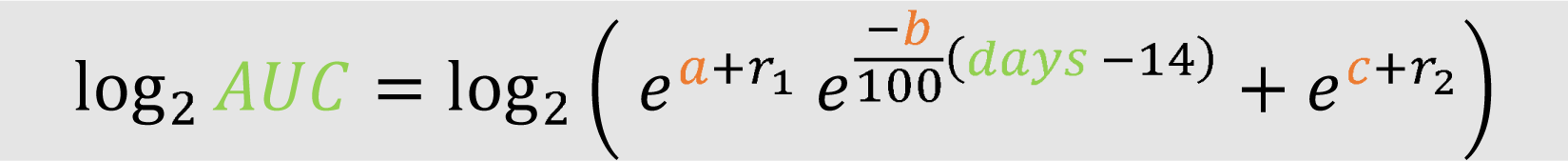

All models were generated using the NLMIXED procedure in SAS9.4. This procedure is capable of fitting arbitrary nonlinear functions including both fixed and random effects. The specific model structure used in this case is shown in **Supplemental Figure 3B**. A two-phase fitting procedure was used. First, a simplified model without random effects was run from a naïve starting point, and the fitted parameter values were used as the initial values for the mixed effects model. In order to account for underlying variability across participants that was not captured by the fixed effects, a random effect was added to each component for the final mixed effect model. In addition to allowing for variability in the overall magnitude of the antibody response, this approach captures observed variability in the kinetics of the response (fold-reduction relative to peak varies among participants). The model was fit independently to datasets for participants with and without pre-vaccine SARS-CoV-2 infection (hybrid immunity) using a feature in SAS to build stratified models. The same model framework was used for both post-vaccine and post-boost models. Data from a post-boost model fit to the combined dataset without accounting for pre-vaccine immune status is also presented, reflecting a close correspondence in post-boost kinetics in both stratified groups.

The impact of demographic factors was assessed in the stratified post-vaccine and post-boost models. In all cases, these factors were assessed as a multiplicative factor across the entire time course studied (manifesting on graphs with log-transformed y axes as a y shift). All demographically informed models were constructed iteratively, beginning with a model including all factors considered (age, gender, race, ethnicity, and primary vaccine type). After each model fitting run, the factor with the highest assessed p value was removed if that p value did not meet a relaxed threshold for statistical significance (p < 0.1). The model was then re-run with the reduced set of fixed effects and finalized when all factors hit this relaxed threshold. This stepwise subtractive modeling practice helps to account for confounding factors and avoids double controlling for effects that alter the result through the same mechanism. Results from the model fitting in SAS were exported to csvs for visualization in Python. SAS code for the model fitting will be made available in a public GitHub repository.

### Supplemental Tables

**Supplemental Table 1:**
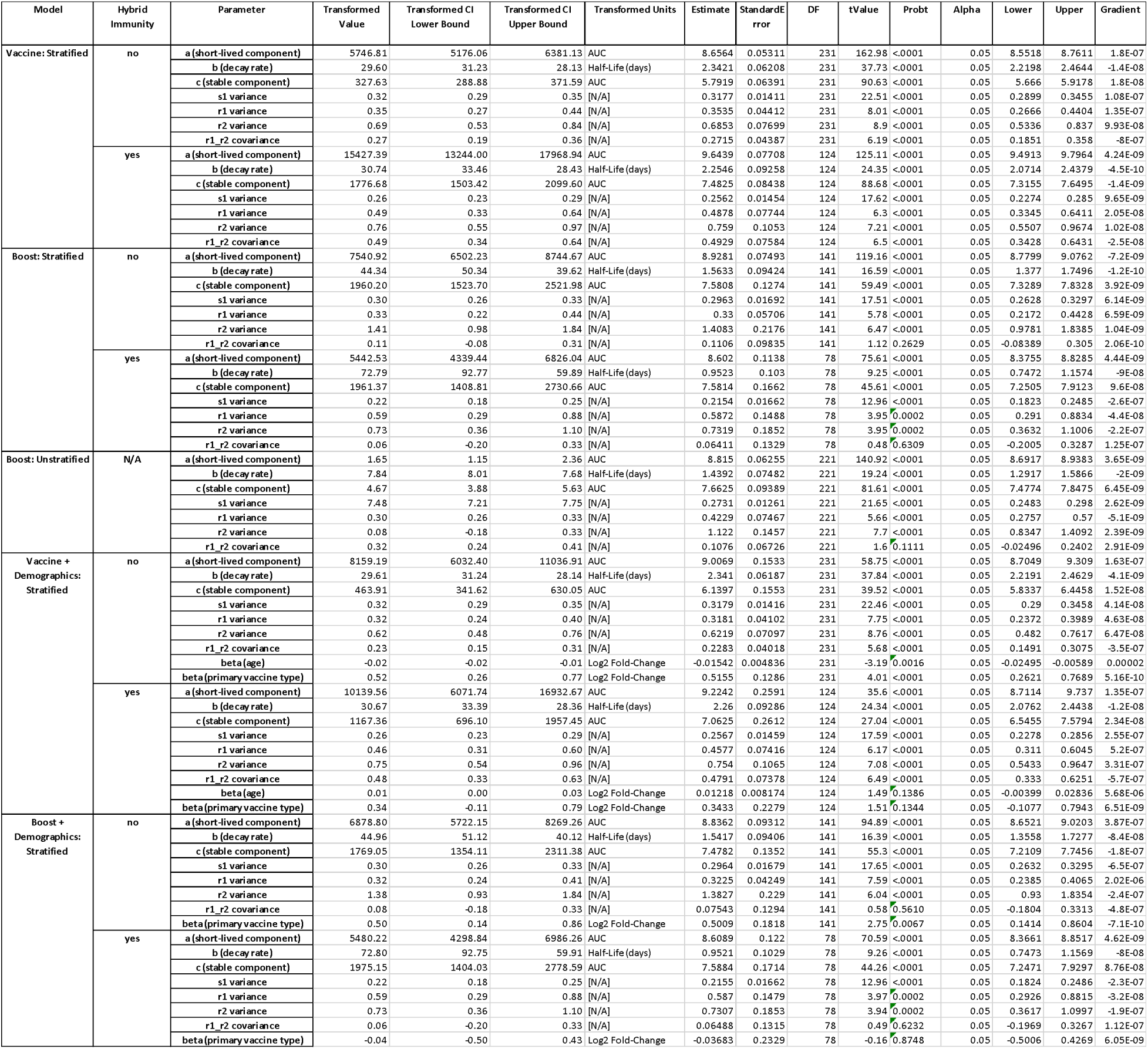
Model development and validations.

**Supplemental Table 2:**
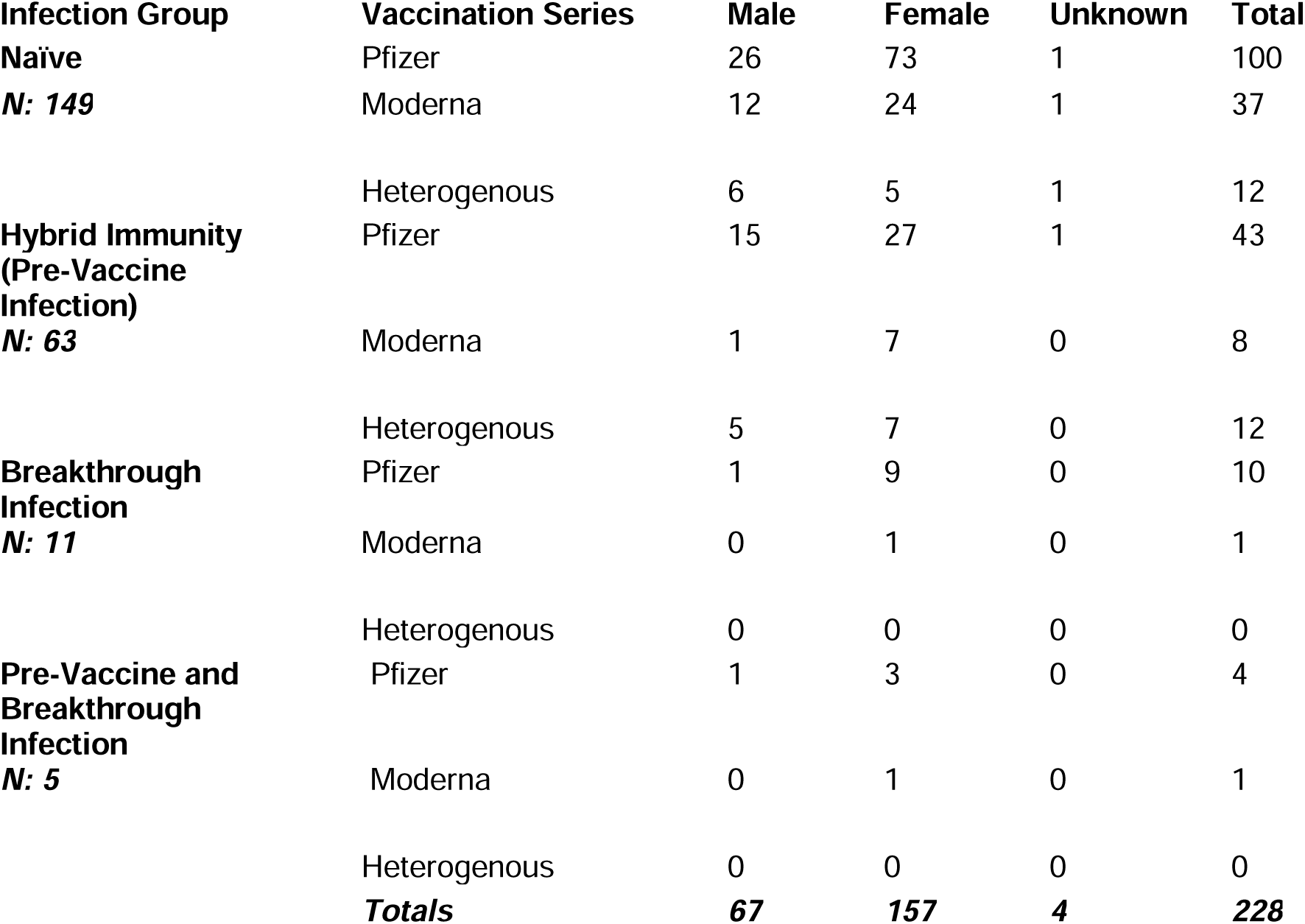
Overview of the immune histories (infection, vaccination) for the 228 PARIS participants who completed all three vaccine reactogenicity surveys.

**Supplemental Table 3:** Vaccine side effects divided by vaccine doses and immunity groups.

|  | Serostatus by Vaccine Type |  |  |  | Total by Vaccine Type |  | Total by Serostatus |  |
| --- | --- | --- | --- | --- | --- | --- | --- | --- |
|  | Naïve (N=160) |  | Hybrid Immunity (N=68) |  | Total (N=228) |  | Total (N=228) |  |
|  | Moderna | Pfizer | Moderna | Pfizer | Moderna | Pfizer | Naïve | Hybrid Immunity |
|  | N=42 (26%) | N=118 (74%) | N=15 (22%) | N=53 (78%) | N=57 (25%) | N=171 (75%) | N=160 (70%) | N=68 (30%) |
| **Local Symptoms** |  |  |  |  |  |  |  |  |
| Pain | 33 (79%) | 86 (73%) | 14 (93%) | 41 (77%) | 47 (82%) | 127 (74%) | 119 (74%) | 55 (81%) |
| Erythema | 7 (17%) | 14 (12%) | 3 (20%) | 5 (9%) | 10 (18%) | 19 (11%) | 21 (13%) | 8 (12%) |
| Induration | 11 (26%) | 17 (14%) | 7 (47%) | 7 (13%) | 18 (32%) | 24 (14%) | 28 (18%) | 14 (21%) |
| Lymphadenopathy | 4 (10%) | 23 (19%) | 1 (7%) | 4 (8%) | 5 (8%) | 27 (16%) | 27 (17%) | 5 (7%) |
| Other | 1 (2%) | 6 (5%) | 4 (27%) | 1 (2%) | 5 (8%) | 7 (4%) | 7 (4%) | 5 (7%) |
| Any Local | 33 (79%) | 90 (76%) | 14 (93%) | 42 (79%) | 47 (82%) | 132 (77%) | 123 (77%) | 56 (82%) |
| **Systemic Symptoms** |  |  |  |  |  |  |  |  |
| Fatigue | 24 (57%) | 80 (68%) | 13 (87%) | 31 (58%) | 37 (65%) | 111 (65%) | 104 (65%) | 44 (65%) |
| Headache | 27 (64%) | 56 (47%) | 9 (60%) | 28 (53%) | 36 (63%) | 81 (47%) | 83 (52%) | 34 (50%) |
| Myalgia | 19 (45%) | 56 (47%) | 10 (67%) | 32 (60%) | 29 (51%) | 77 (45%) | 75 (47%) | 31 (46%) |
| Chills | 17 (40%) | 43 (36%) | 11 (73%) | 20 (38%) | 28 (49%) | 63 (37%) | 60 (37%) | 31 (46%) |
| Arthralgia | 14 (33%) | 19 (16%) | 7 (47%) | 7 (13%) | 21 (37%) | 26 (15%) | 33 (21%) | 14 (21%) |
| Fever | 19 (45%) | 36 (31%) | 9 (60%) | 11 (21%) | 28 (49%) | 47 (27%) | 55 (34%) | 20 (29%) |
| Nausea/Emesis | 7 (17%) | 11 (9%) | 2 (13%) | 7 (13%) | 9 (16%) | 18 (11%) | 54 (34%) | 36 (53%) |
| Pharyngitis | 4 (10%) | 11 (9%) | 1 (7%) | 3 (6%) | 5 (8%) | 14 (8%) | 15 (9%) | 4 (6%) |
| Other | 6 (14%) | 5 (4%) | 0 (0%) | 3 (6%) | 6 (11%) | 8 (5%) | 11 (7%) | 3 (4%) |
| Any Systemic | 33 (79%) | 88 (74%) | 13 (87%) | 40 (75%) | 46 (81%) | 128 (75%) | 121 (76%) | 53 (78%) |

### Supplemental Figures

**Supplemental Figure S1:**
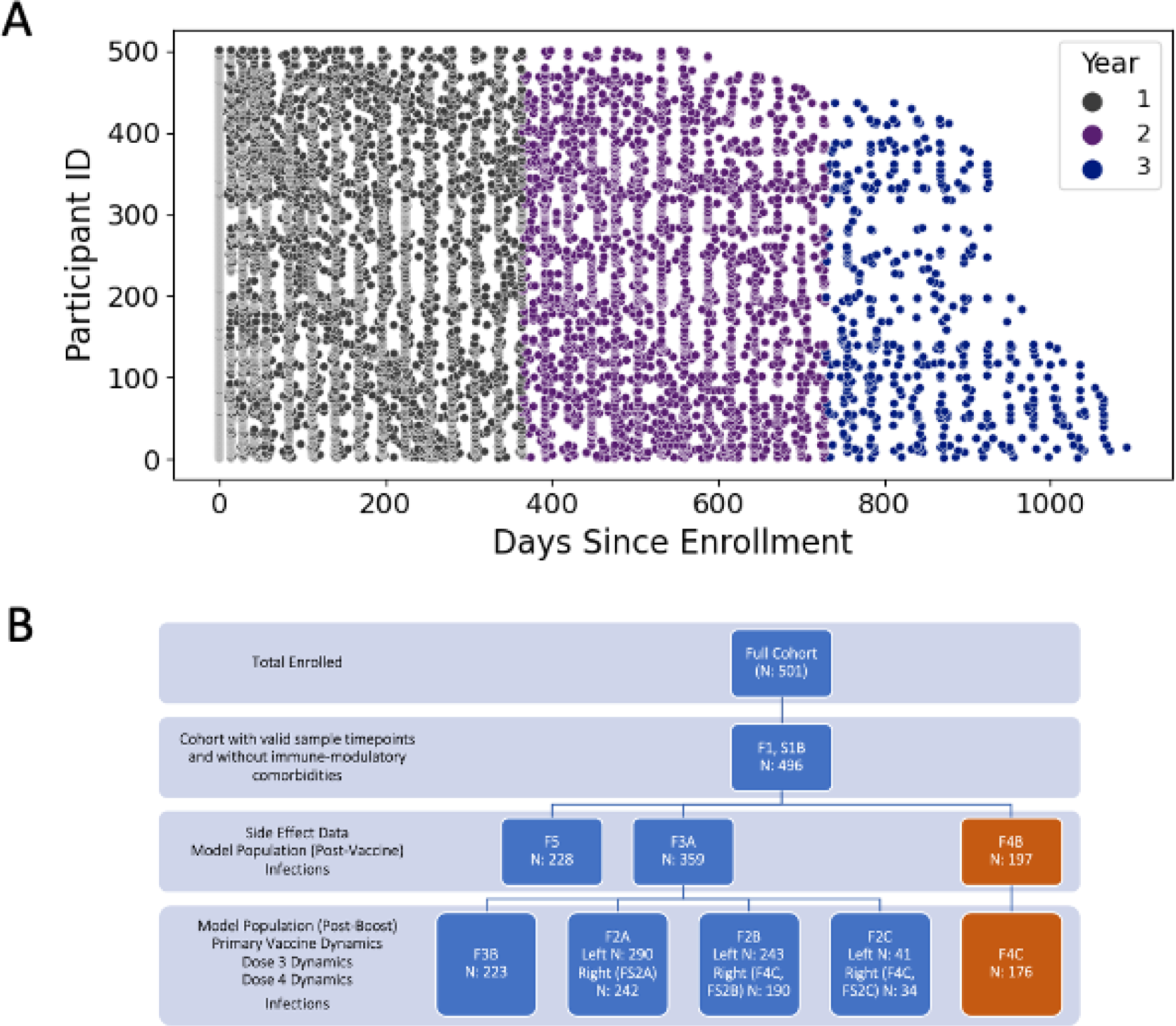
A: Study visits from each of the 496 PARIS participants are shown colored by year of study follow-up. A total of 8,041 samples were collected over the three years of the study. B: Flow chart illustrating the data selection from PARIS study participants for the specific analyses performed. Figures in which a given dataset or subset thereof are shown are listed. Datasets based on infection data are shown in dark orange.

**Supplemental Figure S2:**
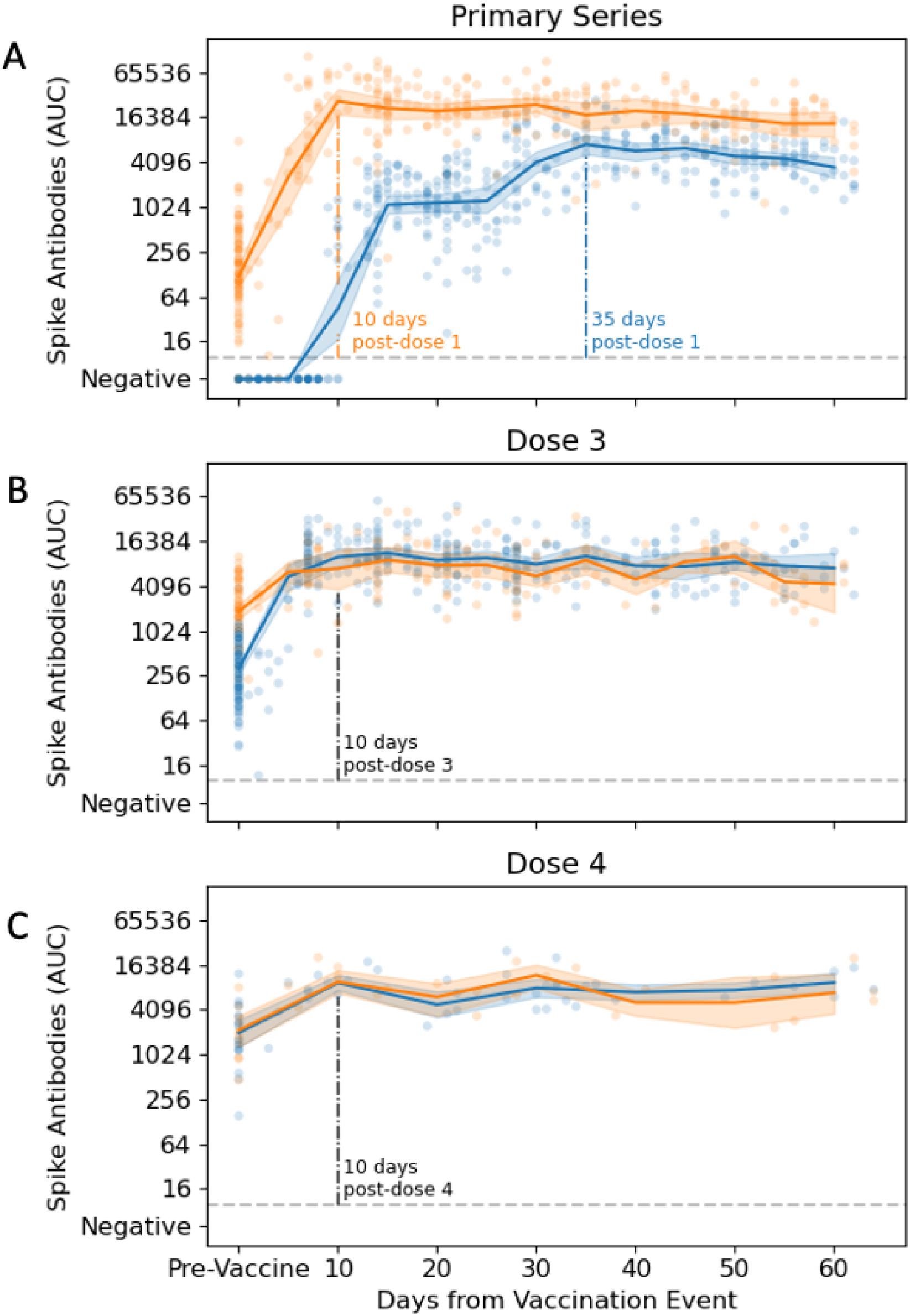
Antibody kinetics after vaccination. Early timepoints post-vaccine. Participants with hybrid immunity due to pre-vaccine SARS-CoV-2 infection are shown in orange, and those without are shown in blue. Participants with breakthrough infections are excluded. Lines connect geometric means for 5-day bins (10-day bins post-dose 4), with confidence intervals constructed by bootstrapping. A: Longitudinal antibody data collected within 40 days after vaccination. Data from participants with (124 participants, 375 samples) and without (166 participants, 528 samples) pre-existing immunity are shown. Approximate peak timepoints for each group are indicated by dashed lines. B: Longitudinal antibody data collected within 40 days after the third vaccine dose. Data for participants with (59 participants, 151 samples) and without (157 participants, 448 samples) prior SARS-CoV-2 infection. C: Longitudinal antibody data collected within 40 days after the 4^th^ vaccine for participants with (12 participants, 28 samples) and without (25 participants, 57 samples) prior SARS-CoV-2 infection

**Supplemental Figure S3:**
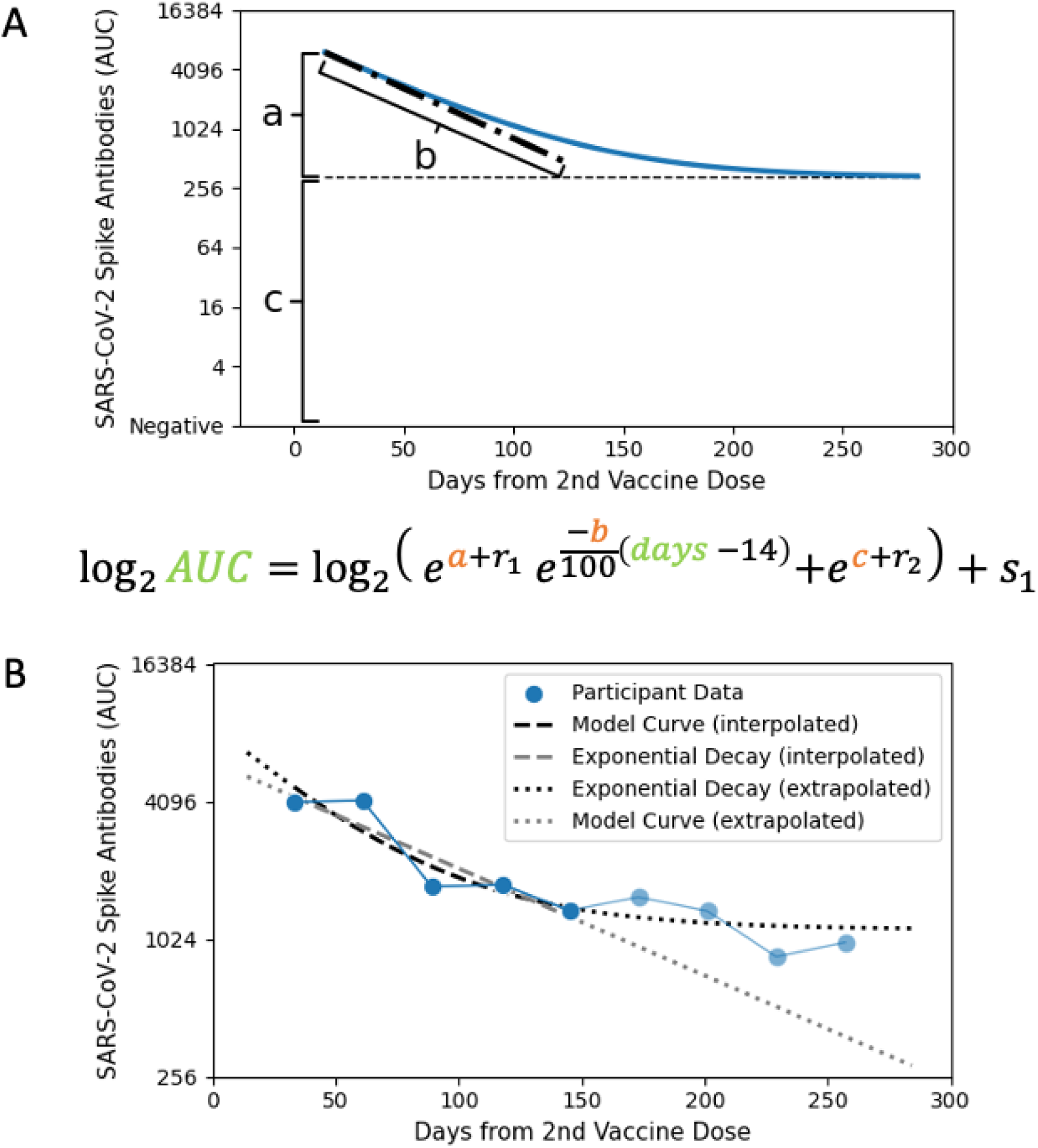
PARIS model development. A: The three parameters of the vaccine are illustrated here, with ‘c’ representing the “steady-state” antibody level, ‘a’ representing the magnitude of the fast-decaying component, and ‘b’ representing the rate of that decay. B: Comparison of a linear fit (grey) and the PARIS model fit (black) to the first five months of data for one representative study participant, with extrapolation from each model plotted against additional timepoints censored from model training.

**Supplemental Figure S4:**
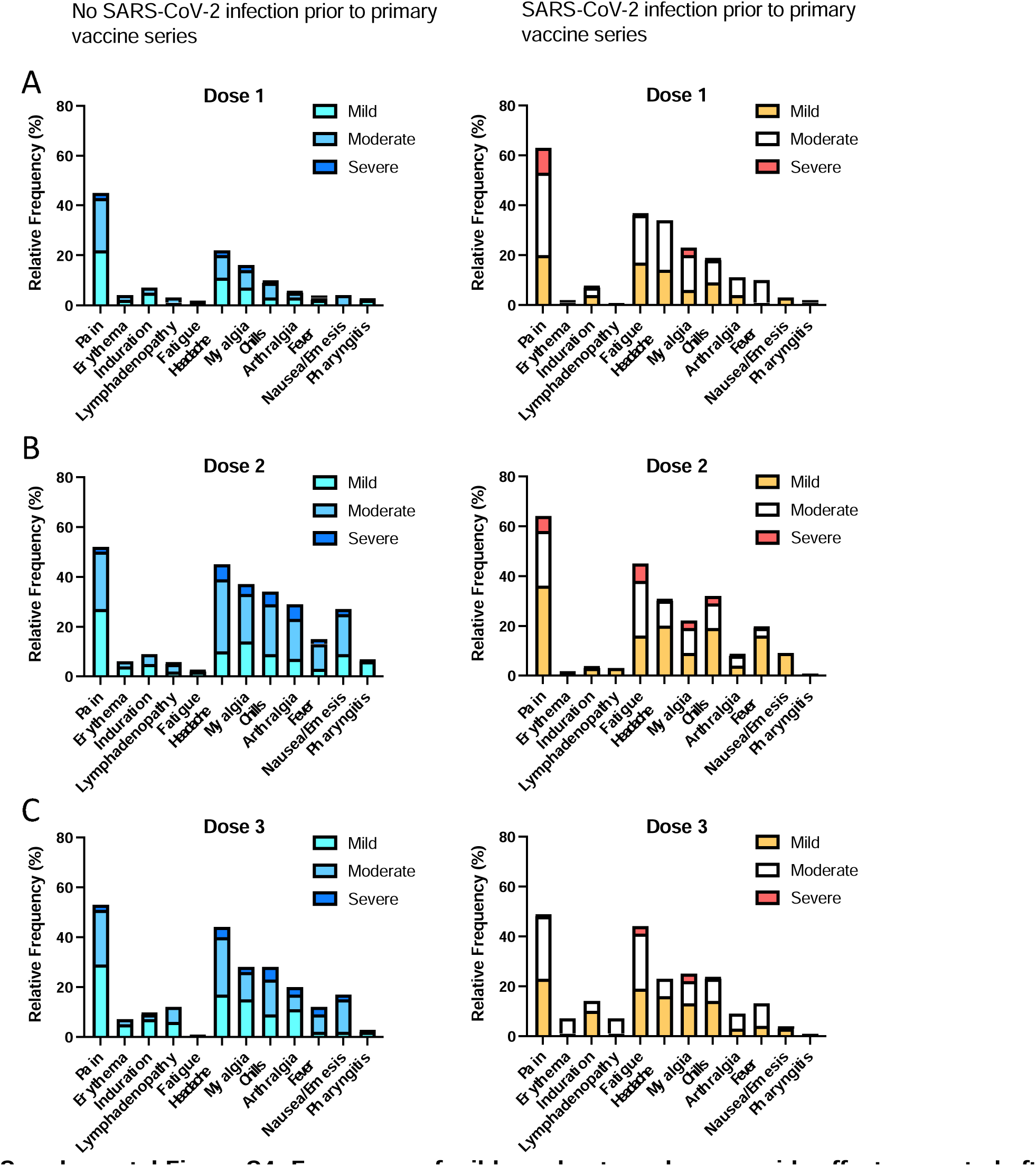
Frequency of mild, moderate and severe side effects reported after SARS-CoV-2 vaccine doses 1, 2 and 3. Local and systemic side effects were recorded after the first (**A**), second (**B**) and third vaccine (**C**) doses in PARIS participants with (right, yellow) and without (left, blue) pre-existing immunity. 230 participants completed all three surveys. Symptom severity was self-reported and classified as mild, moderate, or severe based on description and duration.

## References

1 Carreño, J. M. et al. Activity of convalescent and vaccine serum against SARS-CoV-2 Omicron. Nature (2021). 10.1038/s41586-022-04399-5

2 Menegale, F. et al. Evaluation of Waning of SARS-CoV-2 Vaccine-Induced Immunity: A Systematic Review and Meta-analysis. JAMA Netw Open 6, e2310650 (2023). 10.1001/jamanetworkopen.2023.10650

3 Zaeck, L. M., GeurtsvanKessel, C. H. & de Vries, R. D. COVID-19 vaccine effectiveness and evolving variants: understanding the immunological footprint. Lancet Respir Med 11, 395–396 (2023). 10.1016/S2213-2600(23)00140-6

4 DeGrace, M. M. et al. Defining the risk of SARS-CoV-2 variants on immune protection. Nature 605, 640–652 (2022). 10.1038/s41586-022-04690-5

5 Feikin, D. R. et al. Duration of effectiveness of vaccines against SARS-CoV-2 infection and COVID-19 disease: results of a systematic review and meta-regression. Lancet 399, 924–944 (2022). 10.1016/S0140-6736(22)00152-0

6 Gonzalez-Reiche, A. S. et al. Introductions and early spread of SARS-CoV-2 in the New York City area. Science 369, 297–301 (2020). 10.1126/science.abc1917

7. Wajnberg, A., et al. Humoral response and PCR positivity in patients with COVID-19 in the New York City region, USA: an observational study. Lancet Microbe 1, e283-e289 (2020). 10.1016/s2666-5247(20)30120-8

8 Stadlbauer, D. et al. Repeated cross-sectional sero-monitoring of SARS-CoV-2 in New York City. Nature 590, 146–150 (2021). 10.1038/s41586-020-2912-6

9 Amanat, F. et al. A serological assay to detect SARS-CoV-2 seroconversion in humans. Nat Med 26, 1033–1036 (2020). 10.1038/s41591-020-0913-5

10 Simon, V. et al. PARIS and SPARTA: Finding the Achilles’ Heel of SARS-CoV-2. mSphere 7, e0017922 (2022). 10.1128/msphere.00179-22

11 Kubale, J. et al. SARS-CoV-2 Spike-Binding Antibody Longevity and Protection from Reinfection with Antigenically Similar SARS-CoV-2 Variants. mBio 13, e0178422 (2022). 10.1128/mbio.01784-22

12 Krammer, F. et al. Antibody Responses in Seropositive Persons after a Single Dose of SARS-CoV-2 mRNA Vaccine. N Engl J Med 384, 1372–1374 (2021). 10.1056/NEJMc2101667

13 Krammer, F. The role of vaccines in the COVID-19 pandemic: what have we learned? Semin Immunopathol (2023). 10.1007/s00281-023-00996-2

14 Akkaya, M., Kwak, K. & Pierce, S. K. B cell memory: building two walls of protection against pathogens. Nat Rev Immunol 20, 229–238 (2020). 10.1038/s41577-019-0244-2

15 Wrammert, J. et al. Rapid cloning of high-affinity human monoclonal antibodies against influenza virus. Nature 453, 667–671 (2008). 10.1038/nature06890

16 Ellebedy, A. H. et al. Defining antigen-specific plasmablast and memory B cell subsets in human blood after viral infection or vaccination. Nat Immunol 17, 1226–1234 (2016). 10.1038/ni.3533

17 Mankarious, S. et al. The half-lives of IgG subclasses and specific antibodies in patients with primary immunodeficiency who are receiving intravenously administered immunoglobulin. J Lab Clin Med 112, 634–640 (1988).

18 Turner, J. S. et al. SARS-CoV-2 infection induces long-lived bone marrow plasma cells in humans. Nature 595, 421–425 (2021). 10.1038/s41586-021-03647-4

19 Ioannou, G. N., Locke, E. R., Green, P. K. & Berry, K. Comparison of Moderna versus Pfizer-BioNTech COVID-19 vaccine outcomes: A target trial emulation study in the U.S. Veterans Affairs healthcare system. EClinicalMedicine 45, 101326 (2022). 10.1016/j.eclinm.2022.101326

20 Bajema, K. L. et al. Comparative Effectiveness and Antibody Responses to Moderna and Pfizer-BioNTech COVID-19 Vaccines among Hospitalized Veterans - Five Veterans Affairs Medical Centers, United States, February 1-September 30, 2021. MMWR Morb Mortal Wkly Rep 70, 1700–1705 (2021). 10.15585/mmwr.mm7049a2

21 Atmar, R. L. et al. Homologous and Heterologous Covid-19 Booster Vaccinations. N Engl J Med 386, 1046–1057 (2022). 10.1056/NEJMoa2116414

22 Sano, K. et al. SARS-CoV-2 vaccination induces mucosal antibody responses in previously infected individuals. Nat Commun 13, 5135 (2022). 10.1038/s41467-022-32389-8

23 Sheikh-Mohamed, S. et al. Systemic and mucosal IgA responses are variably induced in response to SARS-CoV-2 mRNA vaccination and are associated with protection against subsequent infection. Mucosal Immunol 15, 799–808 (2022). 10.1038/s41385-022-00511-0

24 Amanat, F. et al. SARS-CoV-2 mRNA vaccination induces functionally diverse antibodies to NTD, RBD, and S2. Cell 184, 3936–3948.e3910 (2021). 10.1016/j.cell.2021.06.005

25 Singh, G. et al. Binding and avidity signatures of polyclonal sera from individuals with different exposure histories to SARS-CoV-2 infection, vaccination, and Omicron breakthrough infections. J Infect Dis (2023). 10.1093/infdis/jiad116

26 Vikström, L. et al. Vaccine-induced correlate of protection against fatal COVID-19 in older and frail adults during waves of neutralization-resistant variants of concern: an observational study. Lancet Reg Health Eur 30, 100646 (2023). 10.1016/j.lanepe.2023.100646

27 Alsoussi, W. B. et al. SARS-CoV-2 Omicron boosting induces de novo B cell response in humans. Nature (2023). 10.1038/s41586-023-06025-4

28 Carreño, J. M., Singh, G., Simon, V., Krammer, F. & group, P. s. Bivalent COVID-19 booster vaccines and the absence of BA.5-specific antibodies. Lancet Microbe 4, e569 (2023). 10.1016/S2666-5247(23)00118-0

29 Fonville, J. M. et al. Antibody landscapes after influenza virus infection or vaccination. Science 346, 996–1000 (2014). 10.1126/science.1256427

30 Polack, F. P. et al. Safety and Efficacy of the BNT162b2 mRNA Covid-19 Vaccine. N Engl J Med (2020). 10.1056/NEJMoa2034577

31 Baden, L. R. et al. Efficacy and Safety of the mRNA-1273 SARS-CoV-2 Vaccine. N Engl J Med (2020). 10.1056/NEJMoa2035389

32 Stadlbauer, D. et al. SARS-CoV-2 Seroconversion in Humans: A Detailed Protocol for a Serological Assay, Antigen Production, and Test Setup. Curr Protoc Microbiol 57, e100 (2020). 10.1002/cpmc.100

